# Microtubule targeting agents influence the clinical benefit of immune response in early breast cancer

**DOI:** 10.1101/2024.03.09.24304017

**Authors:** Vinu Jose, David Venet, Françoise Rothé, Samira Majjaj, Delphine Vincent, Laurence Buisseret, Roberto Salgado, Nicolas Sirtaine, Stefan Michiels, Sherene Loi, Heikki Joensuu, Christos Sotiriou

**Author notes:** Corresponding author Christos Sotiriou, JC Heuson Breast Cancer Translational Research Laboratory (BCTL), Institut Jules Bordet - Université Libre de Bruxelles, Rue Meylemeersch 90, Brussels 1070, Belgium. Conflict of interest: The authors declare the following financial interests/personal relationships which may be considered as potential competing interests: Roberto Salgado reports non-financial support from Merck and Bristol Myers Squibb (BMS), research support from Merck, Puma Biotechnology and Roche, and personal fees from Roche, BMS and Exact Sciences for advisory boards. Stefan Michiels reports fees outside the scope of the submitted work for statistical advice to IDDI, Amaris, Roche, and data and safety monitoring member of clinical trials: IQVIA, Sensorion, Biophytis, Servier, Yuhan. Sherene Loi receives research funding to her institution from Novartis, Bristol Meyers Squibb, Merck, Puma Biotechnology, Eli Lilly, Nektar Therapeutics, Astra Zeneca, Roche-Genentech and Seattle Genetics. She has acted as consultant (not compensated) to Seattle Genetics, Novartis, Bristol Meyers Squibb, Merck, AstraZeneca, Eli Lilly, Pfizer, Gilead Therapeutics and Roche-Genentech. She has acted as consultant (paid to her institution) to Aduro Biotech, Novartis, GlaxoSmithKline, Roche-Genentech, Astra Zeneca, Silverback Therapeutics, G1 Therapeutics, PUMA Biotechnologies, Pfizer, Gilead Therapeutics, Seattle Genetics, Daiichi Sankyo, Merck, Amunix, Tallac Therapeutics, Eli Lilly and Bristol Meyers Squibb. Heikki Joensuu has had a leadership role in Orion Pharma, Neutron Therapeutics, and Sartar Therapeutics, has a consulting/advisory role in Orion Pharma and Neutron Therapeutics, has received honoraria for scientific meetings from Deciphera Pharmaceuticals, and has stock ownership in Orion Pharma and Sartar Therapeutics. Christos Sotiriou has received research grants from Astellas, Cepheid, Vertex, Seattle Genetics, Puma, Amgen, and Merck & has support for attending meetings and/or travel from Roche, Genentech, and received payment or honoraria for lectures, presentations, speakers bureaus, manuscript writing or educational events from Eisai, Prime Oncology, Teva, and Exact Sciences.

## Abstract

**PURPOSE:** Immune response to tumors is associated with clinical benefits in breast cancer. Preclinically, disruption of microtubule dynamics affect the functionality of immune cells. We investigate the impact of microtubule targeting agents (MTA) on the clinical benefit of immune response in early breast cancer.

**METHODS:** We used the gene expression dataset associated with the randomized FinHER adjuvant phase III trial, which compared Docetaxel (stabilizing MTA) to Vinorelbine (destabilizing MTA), and an integrated non-randomized GEO neoadjuvant dataset with regimens containing stabilizing MTA or without any MTA. Cox/logistic interaction models assessed the interaction between MTAs and immune response on clinical benefit. Immune response was measured using histopathology (TIL-H&E), gene module scores, and immune cell-type estimation methods.

**RESULTS:** MTA and immune responses interact significantly in breast cancer, particularly in TNBC, affecting patient survival. In the randomized FinHER adjuvant TNBC setting, a unit increase in interferon score is associated with a death hazard-ratio (HR) of 10.97 (95% confidence interval, 0.79 to 151.78) in the Docetaxel arm (n=60), and a death HR of 0.16 (0.03 to 0.97) in the Vinorelbine arm (n=60), P-interaction = 0.008 (FDR-adjusted, 0.039). In the non-randomized neoadjuvant TNBC setting, a unit increase in interferon score is associated with a pathological-complete-response (pCR) odds-ratio (OR) of 1.3 (0.6 to 3.1) in stabilizing MTA regimens (n=293), and a pCR OR of 46.8 (3.9 to 557.7) in non-MTA regimens (n=83), P-interaction = 0.004 (FDR-adjusted, 0.032).

**CONCLUSION:** MTAs influence the clinical benefit of immune response in breast cancer. However, the limited sample size warrants additional analyses.

**Translational relevance:** Creating combination regimens with immune system stimulation, such as immunotherapy, requires classification of cancer therapies by their effects on immune cells. The finding that microtubule-destabilizing agents respond better to immunogenic TNBCs than stabilizing agents (taxanes), and vice-versa, has different implications. Firstly, destabilizing agents, currently recommended in metastatic settings, can be brought into early settings for immunogenic TNBCs while limiting stabilizing agents to non-immunogenic tumors. Secondly, stabilizing agents may be more effective as backbone therapy for immunotherapy in non-immunogenic tumors than destabilizing agents and vice-versa. Furthermore, the potential use of destabilizing agents as checkpoint inhibitors in immunogenic TNBC is warranted from the present non-immunotherapy dataset. Finally, since routine evaluation of immune response is recommended from tumor biopsies, the heterogeneity observed between TIL counts from histopathology and gene signatures of immune response calls for additional research into the objectivity of different measures of immune response.

## Introduction

In breast cancer (BC), many studies reported the prognostic and predictive value of host immune response to tumor (immune response) (1–3), in which immune response was measured as the percentage of tumor-infiltrating lymphocytes (TIL) by a pathologist using morphological evaluation of hematoxylin-and-eosin (H&E) stained formalin-fixed-paraffin-embedded (FFPE) sections (4). Interestingly, preclinical studies reported the potential impact of microtubule targeting agents (MTA) on immune cell’s functionality (5–7). For instance, Franchini et al. demonstrated T-cell’s dependence on stable microtubules for its stress-granule-dependent inhibitory checkpoint translation (6). Stress granule (SG) formation is a cell mechanism to avoid self-destruction/apoptosis during cellular stress (8). Tumor cells exploit this intrinsic cellular mechanism for survival during cellular stress, such as radiation/chemo/targeted therapies, and hence for treatment resistance (8). Preclinical studies have shown the differential roles of stabilizing and destabilizing MTAs on SG formation and maturation (9,10), which may impact cell physiology differently, including immune cells (11–13). Given that microtubule-targeting agents (MTAs) are used routinely in clinics for multiple cancer types (14), it is intriguing how they interfere with the clinical relevance of immune responses.

The gene expression dataset of the Finland Herceptin (FinHER) adjuvant trial (15,16), comparing the stabilizing MTA, Docetaxel, to the destabilizing MTA, Vinorelbine, provides an ideal setting to investigate the impact of MTAs on the clinical relevance of immune response. Using the FinHER gene expression dataset (17) and an integrated neoadjuvant dataset with detailed characterization of treatment regimens (18), the present study demonstrates the interaction between MTAs and immune response on clinical benefit in the early adjuvant/neoadjuvant breast cancer settings.

## Materials and methods

### FinHER adjuvant dataset

The FinHER phase III clinical trial patients were randomized to receive Docetaxel (stabilizing MTA) or Vinorelbine (destabilizing MTA) chemotherapy (doi.org/10.1186/ISRCTN76560285) (15,16). In addition, all patients received 5-Fluorouracil, Epirubicin, and Cyclophosphamide (FEC) chemotherapy; human-epidermal-growth-factor-receptor-2 (HER2) amplified BC (HER2+BC) patients received trastuzumab or no HER2 therapy based on random allocation; and hormone-receptor (HR) positive BC (HR+BC – esterogen-receptor (ER) or progesteron-receptor (PR) positive BC) patients received hormone therapy. Pre-treatment breast cancer resection specimens were collected from advanced node-positive or high-risk node-negative breast cancer patients enrolled in the trial. Gene expression data were derived from FFPE preserved triple-negative BC (TNBC – ER, PR, and HER2 negative BC) and HER2+BC specimens. Gene expression profiling from degraded FFPE RNA and FFPE specific data processing are detailed elsewhere (17). After sample-probeset filtering, 90% samples (#300) and 13% probesets (#6207 probesets, #3350 genes) remained. The MAS normalized, sample-probeset filtered, and experimental batch effect corrected FFPE gene expression dataset was extracted from Gene-expression-omnibus (GEO, Accession: GSE47994). Stromal TIL quantification using histopathology was detailed elsewhere (3).

### Integrated GEO dataset

The integrated dataset containing 3,736 samples from 28 datasets with 9,184 common genes and corrected for dataset effect using the quantile method was extracted from GEO (Accession: GSE205568). The dataset integration details are available in (18). In the integrated GEO dataset, patients were administered with regimens containing either stabilizing MTAs (taxanes) or without any MTAs. The integrated dataset extracted was further filtered according to the criteria described in Supplementary Figure 1 to reduce bias in the analysis. No adjuvant samples remained after sample filtering. The analysis was limited to regimens containing Anthracycline-Antimetabolite-Alkylating (AAA) agents with stabilizing MTAs (taxanes) or without any MTAs to make the neoadjuvant analysis comparable to FinHER (FEC with Docetaxel / Vinorelbine) in terms of the treatment regimen. Data from regimens with destabilizing MTAs were not available. HR+BC patients received no hormone therapy in the neoadjuvant settings. No interaction analysis was possible with HER2+BC as all patients received taxane-containing regimens; hence discarded. Finally, eight neoadjuvant datasets with 988 pre-treatment samples remained for analysis.

### Immune response measurements

Hosts’ immune response to the tumor was measured using histopathology, gene modules, and in-silico immune cell-type estimation. Histopathology and in-silico cell-type estimation methods measure the presence/magnitude of immune infiltrates irrespective of their phenotype/activation state, whereas gene modules measure the active biological process (BP).

### Histopathology

Two pathologists evaluated morphologically the lymphocytes infiltrating the tumor stroma using H&E stained FFPE tumor sections (TIL-H&E) from the FinHER dataset, and its mean is used (3). TIL quantification protocol was in agreement with TIL working group guidelines (4). TIL-H&E counts were available only in the FinHER adjuvant dataset.

### Gene modules

Gene modules representing the BPs associated with TIL (TIL-BP gene modules) are used to measure immune responses.

### De novo gene modules associated with TIL

De novo TIL-BP gene modules were identified from the FinHER dataset by gene ontology over-representation analysis of up/down-regulated genes significantly associated with TIL-H&E (TILsig) using the DAVID bioinformatics tool (19) available online. The algorithm was applied separately to TNBC, HER2+BC, and combined samples. Over-represented BP terms present in all TILsig versions were considered robustly associated with TIL-H&E independent of subtype. TILsig genes from the combined dataset annotated with the robustly over-represented BP terms were grouped to generate TIL-BP gene modules. The upregulated BPs were 1) antigen-processing-and-presentation, 2) immune-response, 3) interferon-gamma-signaling, and 4) innate-immune-response. The down-regulated BPs were 1) extracellular matrix (ECM) organization and 2) cell adhesion. The different TIL-BP gene modules within upregulated or down-regulated modules were merged separately due to shared genes and the high co-correlation structure of their module scores (Supplementary Figure 2). Along with the above two merged TIL-BP gene modules (immune and ECM related), the TILsig derived from combined samples (HER2+BC and TNBC) is also used to measure immune response. Supplementary Figure 3 details the gene module generation workflow. Supplementary Table 1 lists the de novo TIL-BP gene modules.

### Published TIL-localization associated gene modules

Four BPs (immune, fibrosis, interferon, and cholesterol) have been identified as associated with TIL’s (CD8 T-cell) localization into the tumor margin, tumor stroma, or tumor core in TNBC (20). They are

1. Fibrosis signaling in no TIL infiltration or tumor-margin-restricted infiltration,
2. Immune and cholesterol signaling in tumor-stroma-restricted infiltration, and
3. Immune and interferon signaling in tumor core TIL infiltration.

The above cholesterol module, when used independently, cannot be interpreted as associated with TIL and measures general cholesterol signaling from bulk tumor expression profiles. Therefore, its inclusion in the analysis is solely for completeness (Supplementary Material). In addition to the above four TIL-BP gene modules, multiple published gene modules representing immune (n=4), interferon (n=3), cholesterol (n=5), and fibrosis (n=3) signaling were also used to measure immune response. The usage of multiple published gene modules ensures the robustness of the results to dataset effects (transcriptome snapshots) and module generation methods (21). Gene module processing is detailed in Supplementary Material and Supplementary Table 2 lists the original gene modules extracted and details of gene module extraction.

### In-silico immune cell-type estimation

MCPcounter (22) estimated cell types from the FinHER and integrated GEO datasets. A recent benchmarking experiment comparing popular cell type estimation methods showed comparable results of MCPcounter with other methods (23).

### Gene module reliability test

Not all genes in the original modules were present in the FinHER and GEO datasets. Hence, using the independent TCGA and Metabric datasets available from MetaGxData (24), the original gene modules are correlated to module subsets from the FinHER and GEO datasets. Gene module subsets with module size greater than two and Pearson correlation greater than 0.95 in TCGA and Metabric datasets are considered reliable.

Two versions of module subsets are considered; (1) subsetting by common 1,932 genes present in FinHER and GEO datasets (FinGEO module subset) and (2) subsetting by 9,184 genes in the GEO dataset (GEO module subset). The primary analysis used FinGEO module subsets and facilitated a direct comparison of results between the two datasets. The GEO module subsets are used only in the GEO dataset for supplementary analysis. This is to explore the impact of larger module sizes on primary analysis results. Supplementary Table 2 lists the module subsets and gene module reliability-test statistics.

Among the 19 published gene modules considered, 10 (53 %) FinGEO module subsets (representing two immune, four interferon, one cholesterol, and three fibrosis) and 17 (89 %) GEO module subsets (representing five immune, four interferon, four cholesterol, and four fibrosis) were reliable. The reliable gene module subsets representing similar BPs contain common genes among them, and their module scores are highly co-correlated (Supplementary Figure 4-6). Therefore, within each version of the module subset, reliable module subsets corresponding to similar BPs were merged. The analysis considers only these pooled modules (Supplementary Table 2).

Similarly, among the nine MCPcounter marker gene modules tested not all of them were reliable. In the FinGEO (marker) module subsets tested, only fibroblast was reliable, while in the GEO (marker) module subsets, T-cell, B-cell-lineage, monocytic-lineage, myeloid-dendritic, endothelial, and fibroblast were reliable. The analysis considers only MCP counter estimates from the reliable marker module subsets (Supplementary Table 2).

### Statistical analysis

The module scores, computed as the difference between the average up-regulated and down-regulated genes on the log scale (25), are range-scaled with minimum and maximum values of 2.5% and 97.5% quantiles, separately for FinHER and GEO datasets. The interaction between MTAs and immune response on clinical benefit is explored in the adjuvant setting using Cox interaction modeling and Kaplan–Meier analysis, and in the neoadjuvant settings using logistic interaction modeling.

Regression analysis results are reported using forest plots. For all Cox models, the PH assumption is tested using the Schoenfeld residual test. A dataset term is included in all logistic regression models (GEO neoadjuvant dataset) to account for the dataset effect. Further, to indicate the degree of dataset heterogeneity that has been accounted for by the dataset term in the logistic regression models, heterogeneity assessment using dataset level summaries in a meta-analytic fashion was performed with the I^2^ statistic and Cochran’s Q statistic (26). Dataset heterogeneity was considered significant if I^2^ statistic >= 0.4 and Cochran’s Q statistic p-value is <0.1. Gene-module significance is defined as unadjusted p-value <0.05 and false discovery rate (FDR) adjusted p-value <0.1. All analyses were performed with R statistical software using the Rstudio IDE. Supplementary material contains additional details. The R scripts are available on GitHub (github.com/vinujose25/taxane-immune-breast, Version 1.0.1,doi.org/10.5281/zenodo.10800408) and OSF (osf.io/urt9x, doi.org/10.17605/OSF.IO/URT9X).

### Ethics statement

The FinHER study participants provided signed informed consent for the clinical trial and signed consent for carrying out research analyses on their tumor tissue. In addition, the ethics committee of Helsinki University Hospital, Finland, (Permission: HUS 177/13/03/02/2011; www.hus.fi) approved a study protocol to perform gene expression and TIL analyses from the specimens collected during the FinHER study.

## Results

### Dataset characteristics

The processed adjuvant FinHER FFPE dataset has 120 TNBC and 180 HER2+BC gene expression profiles with 3350 reliable genes available for analysis. The processed neoadjuvant GEO dataset has 376 TNBC and 612 HR+BC gene expression profiles with 9184 genes available for analysis. Tables 1 and 2 detail the clinical and pathological characteristics of tumor samples selected for analysis from the FinHER and GEO datasets. Table 3 provides breast cancer subtype-specific chemotherapy regimens and the number of patients treated with each regimen.

**Table 1:** Clinical and pathological sample characteristics of the adjuvant FinHER dataset.

|  |  |  |
| --- | --- | --- |
| Dataset |  | FinHER |
| Sample size |  | 300 |
| Age | <=50 | 133 |
|  | >50 | 167 |
|  | NA | 0 |
| Grade | 1 | 0 |
|  | 2 | 84 |
|  | 3 | 208 |
|  | NA | 8 |
| Node | Neg | 64 |
|  | Pos | 236 |
|  | NA | 0 |
| Size | T0-1 | 77 |
|  | T2 | 193 |
|  | T3 | 29 |
|  | T4 | 0 |
|  | NA | 1 |
| ER | Neg | 213 |
|  | Pos | 87 |
|  | NA | 0 |
| PR | Neg | 236 |
|  | Pos | 64 |
|  | NA | 0 |
| HER2 | Neg | 120 |
|  | Pos | 180 |
|  | NA | 0 |
| Subtype | HR+BC | 0 |
|  | HER2+BC | 180 |
|  | TNBC | 120 |
|  | NA | 0 |
| Chemotherapy | FEC+Docetaxel | 147 |
|  | FEC+Vinorelbine | 153 |
|  | NA | 0 |
| HER2 therapy | Trastuzumab | 89 |
|  | No Trastuzumab | 91 |
|  | NA | 120 |
| Hormone therapy | Yes | 91 |
|  | No | 209 |
|  | NA | 0 |
| Event | DDFS | 72 |
|  | RFS | 80 |
|  | OS | 48 |

**Table 2:** Clinical and pathological sample characteristics of the neoadjuvant GEO dataset. AAA: Anthracycline-Antimetabolite-Alkylating agent, pCR – pathological-complete-response.

|  |  | All | GSE25066 | GSE20194 | GSE20271 | GSE22093 | GSE32646 | GSE23988 | GSE42822 | GSE50948 |
| --- | --- | --- | --- | --- | --- | --- | --- | --- | --- | --- |
| Sample size |  | 988 | 313 | 205 | 152 | 97 | 81 | 61 | 53 | 26 |
| Age | <=50 | 525 | 163 | 105 | 83 | 54 | 40 | 35 | 32 | 13 |
|  | >50 | 462 | 150 | 100 | 69 | 42 | 41 | 26 | 21 | 13 |
|  | NA | 1 | 0 | 0 | 0 | 1 | 0 | 0 | 0 | 0 |
| Grade | 1 | 59 | 16 | 12 | 14 | 3 | 13 | 1 | 0 | 0 |
|  | 2 | 378 | 117 | 91 | 54 | 29 | 54 | 19 | 0 | 14 |
|  | 3 | 476 | 178 | 97 | 58 | 47 | 14 | 37 | 34 | 11 |
|  | NA | 75 | 2 | 5 | 26 | 18 | 0 | 4 | 19 | 1 |
| Node | neg | 289 | 96 | 61 | 49 | 21 | 23 | 21 | 18 | 0 |
|  | pos | 624 | 217 | 144 | 102 | 29 | 58 | 40 | 34 | 0 |
|  | NA | 75 | 0 | 0 | 1 | 47 | 0 | 0 | 1 | 26 |
| Size | T0-1 | 63 | 25 | 18 | 11 | 3 | 4 | 1 | 1 | 0 |
|  | T2 | 504 | 171 | 116 | 64 | 51 | 62 | 20 | 20 | 0 |
|  | T3 | 253 | 77 | 38 | 31 | 26 | 12 | 40 | 29 | 0 |
|  | T4 | 142 | 40 | 33 | 46 | 17 | 3 | 0 | 3 | 0 |
|  | NA | 26 | 0 | 0 | 0 | 0 | 0 | 0 | 0 | 26 |
| ER | neg | 407 | 134 | 71 | 63 | 55 | 26 | 29 | 28 | 1 |
|  | pos | 581 | 179 | 134 | 89 | 42 | 55 | 32 | 25 | 25 |
|  | NA | 0 | 0 | 0 | 0 | 0 | 0 | 0 | 0 | 0 |
| PR | neg | 432 | 166 | 109 | 77 | 0 | 43 | 0 | 28 | 9 |
|  | pos | 398 | 147 | 96 | 75 | 0 | 38 | 0 | 25 | 17 |
|  | NA | 158 | 0 | 0 | 0 | 97 | 0 | 61 | 0 | 0 |
| HER2 | neg | 988 | 313 | 205 | 152 | 97 | 81 | 61 | 53 | 26 |
|  | pos | 0 | 0 | 0 | 0 | 0 | 0 | 0 | 0 | 0 |
|  | NA | 0 | 0 | 0 | 0 | 0 | 0 | 0 | 0 | 0 |
| Subtype | HR | 612 | 194 | 141 | 93 | 42 | 55 | 32 | 29 | 26 |
|  | HER2 | 0 | 0 | 0 | 0 | 0 | 0 | 0 | 0 | 0 |
|  | TN | 376 | 119 | 64 | 59 | 55 | 26 | 29 | 24 | 0 |
|  | NA | 0 | 0 | 0 | 0 | 0 | 0 | 0 | 0 | 0 |
| Arm | AAA | 174 | 0 | 0 | 77 | 97 | 0 | 0 | 0 | 0 |
|  | AAA+Taxane | 814 | 313 | 205 | 75 | 0 | 81 | 61 | 53 | 26 |
|  | NA | 0 | 0 | 0 | 0 | 0 | 0 | 0 | 0 | 0 |
| pCR | No | 783 | 248 | 171 | 133 | 69 | 66 | 41 | 33 | 22 |
|  | Yes | 205 | 65 | 34 | 19 | 28 | 15 | 20 | 20 | 4 |
|  | NA | 0 | 0 | 0 | 0 | 0 | 0 | 0 | 0 | 0 |

**Table 3:** Breast cancer subtype-specific regimen size available for interaction analysis in the FinHER adjuvant and neoadjuvant GEO datasets. TNBC: Triple-negative breast cancer. HER2+BC: HER2 amplified breast cancer. HR+BC: Hormone-receptor-positive breast cancer. FEC: Fluorouracil (Antimetabolite)-Epirubicin (Anthracycline)-Cyclophosphamide (Alkylating agent). AAA: Anthracycline-Antimetabolite-Alkylating agent. ^a^ All hormone-receptor-positive patients received hormone therapy. ^b^ Trastuzumab therapy was randomly assigned, or no HER2 therapy was given. ^c^ No patients received hormone therapy.

| Treatment regimen | HR+BC | HER2+BC | TNBC |
| --- | --- | --- | --- |
| FinHER adjuvant dataset - Randomized |  |  |  |
| FEC + Docetaxel | - | 87<sup>ab</sup> | 60 |
| FEC + Vinorelbine | - | 93<sup>ab</sup> | 60 |
| GEO neoadjuvant dataset - Non-randomized |  |  |  |
| AAA + Taxane | 521<sup>c</sup> | - | 293 |
| AAA | 91<sup>c</sup> | - | 83 |

### MTAs seem to interact with the host immune response in adjuvant and neoadjuvant TNBC

Independent studies reported the clinical response associated with immune response in adjuvant and neoadjuvant TNBC settings (1–3). The present study further demonstrates that MTAs affect the clinical response associated with immune response in TNBC, supporting the preclinical data (Figures 1A, 2, 3A, Supplementary Figures 7-14). In FinHER adjuvant TNBC, a unit increase in interferon score was associated with a death hazard ratio of 10.97 (95% confidence interval, 0.79 to 151.78) in the Docetaxel arm and a death hazard ratio of 0.16 (0.03 to 0.97) in the Vinorelbine arm, P-interaction = 0.008 (FDR-adjusted, 0.039). In addition to the overall survival (OS) data results above, distant disease-free survival (DDFS) and recurrence-free survival (RFS) data give similar results with interferon scores (Figure 1A, Supplementary Figures 7A, and 8A). Further, in GEO neoadjuvant TNBC, a unit increase in interferon score results in a pathological-complete-response (pCR) odds ratio of 1.3 (0.6 to 3.1) in the taxane regimen and a pCR odds ratio of 46.8 (3.9 to 557.7) in the non-MTA regimen, P-interaction = 0.004 (FDR-adjusted, 0.032) (Figure 3A). The non-MTA regimen’s wider confidence interval of odds ratio could be attributed to the small sample size in this regimen (n=83, 22%) compared to the taxane regimen (n=293, 78%) (Table 3). Given that immune or interferon gene module scores are a surrogate for immune response or immune cell infiltration, the above observations imply that stabilized and destabilized microtubules may have a profound differentiating role in immune cells’ function and patient survival.

**Figure 1:**
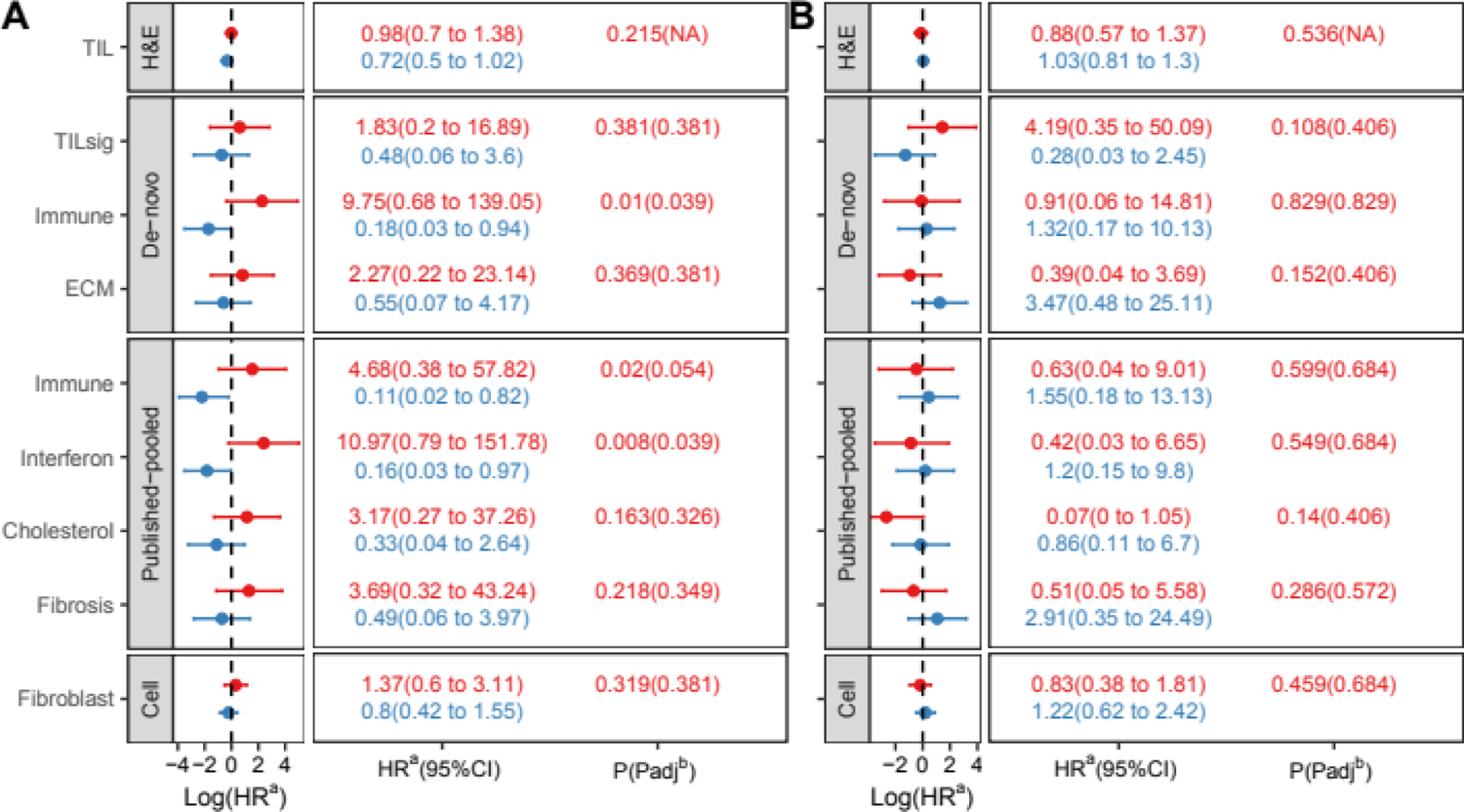
OS interaction between MTAs and immune response measures (TIL-H&E/FinGEO module subsets) in the FinHER adjuvant dataset. (A) TNBC; (B) HER2+BC. A positive log(HR) (i.e. HR>1) indicates a higher death rate, while a negative log(HR) (i.e. HR<1) indicates a lower death rate with an increase in module score in respective treatment regimens. Red represents Docetaxel - stabilizing MTA, and blue represents Vinorelbine - destabilizing MTA. ^a^ Hazard ratio. ^b^ TIL-H&E interaction p-value is not used for FDR correction.

Contrary to TIL-BP gene modules, TIL-H&E did not significantly interact with MTAs in FinHER adjuvant TNBC (P-interaction = 0.215/0.142/0.182 with OS/DDFS/RFS), suggesting potential heterogeneity in immune response measures (Figure 1A, Supplementary Figures 7A and 8A). Notably, Kaplan-Meier analysis revealed a significant clinical benefit with Vinorelbine for high TIL-H&E patients (log-rank P = 0.013/0.005/0.029 for OS/DDFS/RFS) but no response with Docetaxel (log-rank P = 0.54/0.98/0.83 with OS/DDFS/RFS) (Figure 2, Supplementary Figures 9 and 10).

**Figure 2:**
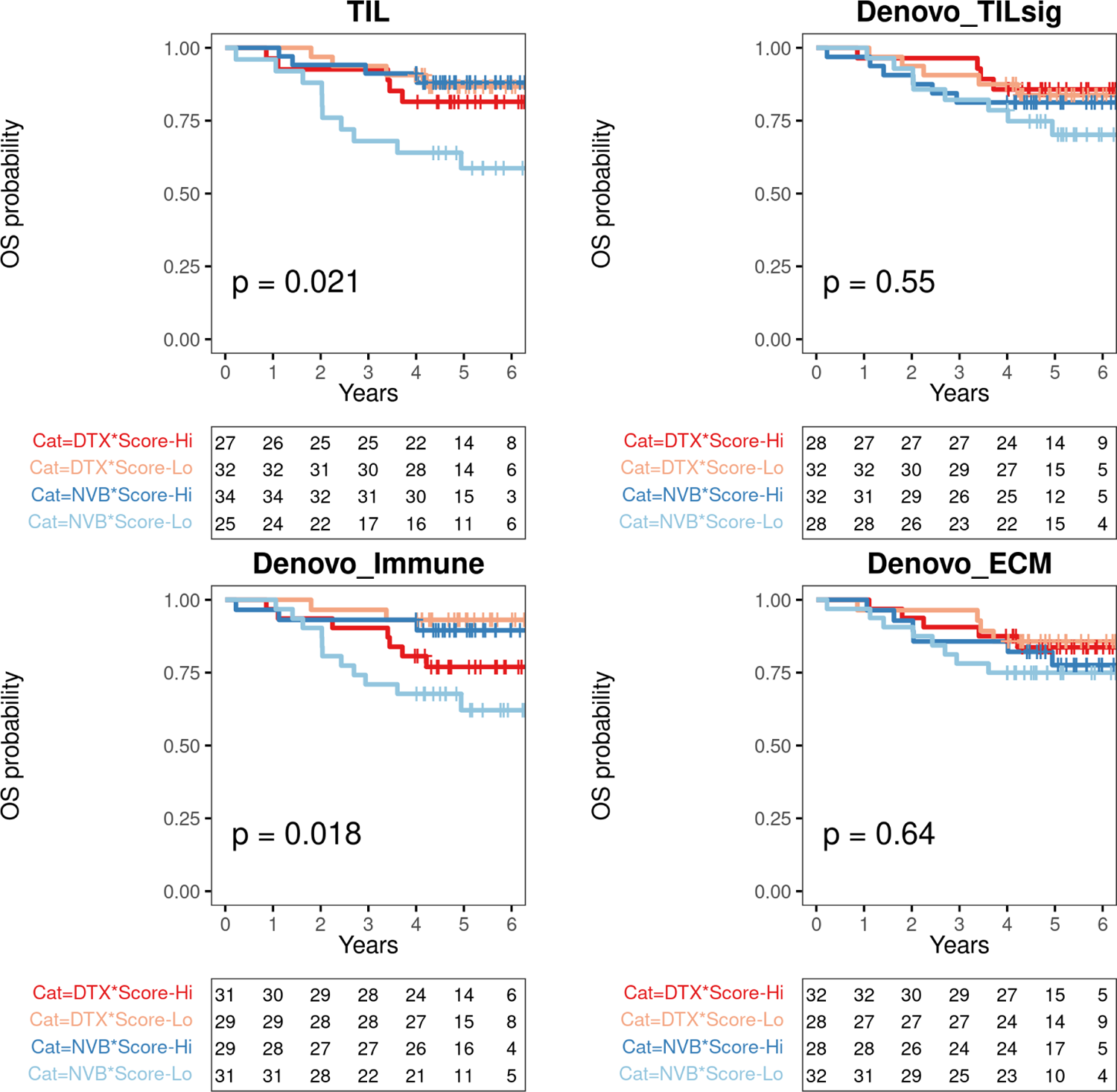
OS Kaplan-Meyer plots reveal the interaction between MTAs and selected immune response measures (TIL-H&E and de novo gene modules) in adjuvant FinHER TNBC. Dark and light red represent Docetaxel with high and low scores respectively. Dark and light blue represent Vinorelbine with high and low scores respectively. High/low classification is based on corresponding medians from TNBC. The median score for TIL (TIL-H&E) in TNBC is 25%. The P-value is computed using the Log-rank test. The table below each Kaplan-Meyer plot shows the number of patients at risk at each time point for each stratum. DTX: Docetaxel - stabilizing MTA. NVB: Vinorelbine - destabilizing MTA.

Further, supporting fibrosis’ association with taxane resistance reported previously (27), increasing fibroblast cell estimates displayed better response from non-MTA regimen than taxane regimen in neoadjuvant TNBC, P-interaction: 0.017 (FDR-adjusted, 0.068) (Figure 3A). A weak interaction trend between MTAs and ECM gene modules is also present in neoadjuvant TNBC (Figures 3A and 14A). However, this phenomenon was not observed in adjuvant TNBC, as the fibrotic tumor tissue is surgically removed before any treatment (Figure 1A).

**Figure 3:**
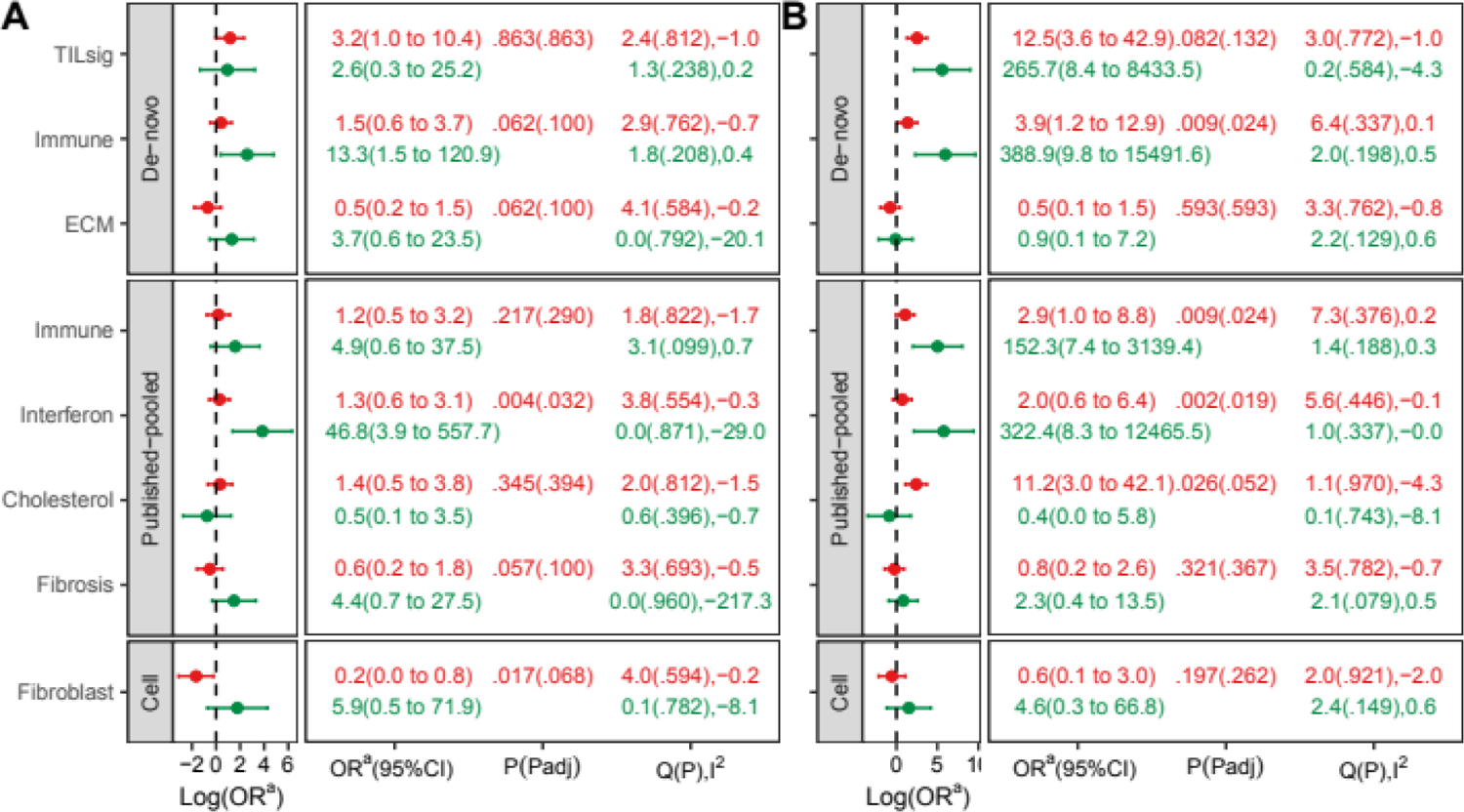
pCR interaction between stabilizing-MTA/no-MTA agent and FinGEO module subsets in the integrated GEO neoadjuvant dataset. (A) TNBC and (B) HR+BC. A positive log(OR) (i.e. OR>1) indicates a higher pCR rate, while a negative log(OR) (i.e. OR<1) indicates a lower pCR rate with an increase in module score in respective treatment regimens. The degree of dataset heterogeneity in each arm is indicated by Cochran’s Q statistic and I^2^ statistic. Q(P),I^2^ represent respectively Cochran’s Q statistic (P-value associated with Q statistic estimated using resampling method) and I^2^ statistic. Red color represents the AAA + Taxane (stabilizing-MTA) regimen, and green color represents the AAA (no-MTA) regimen. ^a^ Odds Ratio.

### MTAs did not interact with host immune response in the adjuvant HER2+BC

No immune response measures significantly interact with MTAs in HER2+BC (Figure 1B, Supplementary Figures 7B, and 8B). It should be noted that the FinHER adjuvant HER2+BC setting had eight treatment regimens, combining Docetaxel/Vinorelbine, Trastuzumab/no-Trastuzumab, and Hormone/no-Hormone therapies, resulting in an approximate sample size of 20 per regimen. Additionally, the clinical relevance of immune responses in adjuvant HER2+BC remains controversial. In HER2+BC patients with anti-HER2 treatment, increased TIL-H&E is associated with improved clinical response (3,28), while in HER2+BC patients without anti-HER2 treatment, TIL-H&E’s prognostic (2) and non-prognostic (29) value has been reported.

### MTAs seem to interact with host immune response in the neoadjuvant HR+BC

Recently, Denkert et al. reported immune response’s clinical relevance in the neoadjuvant HR+BC setting, although not reflected in patient prognosis (1). The present study demonstrates that MTAs could affect immune response’s clinical relevance in HR+BC (Figure 3B and Supplementary Figure 14B). Like adjuvant and neoadjuvant TNBC, a unit increase in interferon score gives a pCR odds ratio of 2.0 (0.6 to 6.4) in taxane regimens and a pCR odds ratio of 322.4 (8.3 to 12465.5) in non-MTA regimens of neoadjuvant HR+BC, P-interaction = 0.002 (FDR-adjusted, 0.019) (Figure 3B). The low sample size of the non-MTA regimen (n=91, 15%) compared to the taxane regimen (n=521, 85%) explains the wider confidence interval of the odds ratio (Table 3).

Further, a higher cholesterol module score is associated with a better response to taxane regimens than non-MTA regimens, P-interaction = 0.026 (FDR-adjusted, 0.052) (Figure 3B). Notably, cholesterol promotes proliferation (30), and proliferating cancers would respond better to regimens containing more therapies simply due to the additive drug effect.

## Discussion

The present retrospective study revealed a potential interaction between MTA and immune response in breast cancer patient survival, particularly in TNBC. In the FinHER adjuvant setting, the destabilizing MTA, Vinorelbine, displayed a survival benefit in immunogenic TNBC, whereas the stabilizing MTA, Docetaxel, displayed potential resistance (Figures 1A and 2, Supplementary Figures 7-13).

A plausible mechanistic explanation for this differential effect can be attributed to the post-transcriptional regulation of immune cells via stress granule dynamics which in turn depends on microtubules (11–13). Stress granule formation in cells is a physiological response to preserve energy for damage repair under stress, such that the cell selectively represses the translation of non-essential gene transcripts to conserve energy for stress-induced damage repair by up-regulating repair protein translation (31). Preclinical studies demonstrated that microtubule stabilizers can augment the formation of large mature stress granules whereas destabilizers could inhibit this (10). Antigen presentation is recognized as a stress signal by T cells.

Hence along with T-cell activation, a stress-granule-based post-transcriptional regulation in T-cells is also activated to fine-tune its response. However, the extent of this post-transcriptional regulation is currently unclear (6,13). One such mechanism is the regulation of immune inhibitory checkpoint translation in CD8, CD4, and gamma-delta T-cells (6). Upon T-cell activation, immune inhibitory checkpoint transcripts, such as PD1 and CTLA4, are formed into stress granules. This step is essential for the spatial segregation of protein synthesis, a level of post-transcriptional regulation wherein polysome assembly and protein synthesis are regulated spatiotemporally (6,11). Microtubule-dependent stress granule formation and transportation seem to play a vital role in this such that inhibitors of stress granules or microtubule dynamics could interfere with this post-transcriptional regulation of immune inhibitory checkpoint protein expression. Among microtubule dynamics inhibitors, destabilizing MTAs are more potent inhibitors of stress-granule-dependent immune inhibitory checkpoint translation in CD8, CD4, and gamma-delta T-cells than stabilizing MTAs (6). Given that CD8-positive and CD4-positive T-cells were the predominant proportion of lymphocytes infiltrating the TNBC (32), the opposing direction of effect in Docetaxel and Vinorelbine according to immunogenicity of TNBC in FinHER dataset, seems to support the microtubule and stress granule dependent post-transcriptional regulation of immune inhibitory checkpoint expression as a mechanism behind the observed differential effect.

Although to a lesser extent, compared to destabilizers, microtubule stabilizers could still impair immune inhibitory checkpoint expression (6). However, this effect in vivo seems negligible, as evidenced by the potential resistance to Docetaxel in FinHER TNBC and the reduced response of neoadjuvant taxane regimens to immunogenic TNBC/HR+BC (Figures 1-3, Supplementary Figures 7-14). Note that stress-granule-based tumor cell resistance due to taxanes (microtubule stabilizers) (8–10) is equally valid for immunogenic and non-immunogenic tumors. Unknown mechanisms may exist behind immunogenic TNBCs’ reduced response to taxanes. One possibility is stress-granule-dependent post-transcriptional regulation of lymphocytes (13,33).

The original FinHER trial reported Docetaxel’s superiority over Vinorelbine (15,16). However, the present study suggests that when tumor immunogenicity is used as a stratifying factor in TNBC, a differential effect is observed between stabilizing and destabilizing agents. Interestingly, this differential effect is observed when gene module scores are used to measure immune response, but not when pathologists’ TIL counts (TIL H&E) are used. Disease management with the less toxic Vinorelbine would be easier and more cost-effective than taxane administration (16,34). Hence, the differential effect observed in the present study has translational relevance, such that Vinorelbine, which is currently limited to advanced metastatic breast cancer settings (35,36), can be brought into the early setting for immunogenic TNBCs and Docetaxel can be limited to non-immunogenic TNBCs. Notably, regimens without MTAs also showed a better response in immunogenic TNBC/HR+BC than taxane-containing regimens in the neoadjuvant setting (Figure 3, Supplementary Figure 14). It is currently unclear whether regimens with microtubule destabilizers are superior to regimens without MTAs in immunogenic tumors.

Recently, ICIs have received accelerated approval from the United States Food and Drug Administration (FDA) for use in TNBC; Pembrolizumab in early settings; Pembrolizumab and Atezolizumab in the advanced metastatic setting (37). However, FDA recommends a chemotherapy backbone for all ICIs, but the optimal chemotherapy backbone to use with these ICIs and their optimum sequencing is currently unknown (37). Further, immunotherapy’s high cost limits its widespread adoption in low-income countries (38). Therefore, the present study’s results have two implications. First, the use of destabilizing MTAs as a potential low-cost ICI version. Second, the use of destabilizing/stabilizing MTAs as an optimal chemotherapy backbone for immunotherapy regimensin immunogenic/non-immunogenic tumors, respectively.

The notion to use destabilizing MTA as an ICI is not novel. In 1990, North and Awwad suggested using Vinblastine as an immunotherapy, observing its ability to relieve suppressed cytotoxicity in CD8 T-cells in a lymphoma model (39).

Additionally, the intracellular mechanism of microtubule agents on T-cells to inhibit immune inhibitory checkpoint expression is complementary to the cell-extrinsic mechanism of ICI agents (6,40). The synergistic effect of anti-PD1 ICI in combination with Vinorelbine (destabilizing agent) and Cyclophosphamide is demonstrated preclinically with 4T1 TNBC syngeneic mouse models (41,42). The above study design lacks a direct comparison between stabilizing and destabilizing agents with or without ICI as the impact of microtubule biology on T-cells was not their prime objective. However, they demonstrated that compared to the taxane-containing anti-PD1 ICI regimen, the regimen with Cyclophosphamide and Vinorelbine has better tumor regression and reduced lung metastasis. When anti-PD1 ICI is added to Cyclophosphamide and Vinorelbine, tumor regression is further improved and lung metastasis is further reduced. Additionally, the authors demonstrated that the efficacy of the Cyclophosphamide and Vinorelbine regimen is mediated through CD8 T-cells. Note that in the above study, the immunogenic Cyclophosphamide is administered before Vinorelbine, suggesting a pre-established immune response before Vinorelbine administration (41). Pre-existing tumor immunogenicity seems essential for the efficacy of destabilizing agents as demonstrated in the present study using Vinorelbine and also by (39) using Vinblastine in lymphoma models. Specifically, Vinblastine’s effectiveness in lymphoma models is dependent on pre-existing CD8 T-cells, such that depletion of CD8 T-cells inhibits tumor regression. Hence Vinorelbine/destabilizing agents would be an ideal chemo backbone for ICI in immunogenic TNBC.

The immunogenicity of taxanes, the ability to elicit an immune response against tumors, is well accepted and is the prime reason for the current combination of taxanes with ICI in TNBC (37,43). In the present study, the superiority of taxanes over vinorelbine in non-immunogenic TNBC could be partially attributed to taxane’s ability to trigger a de novo immune response against tumors. Further, the trend of resistance observed with Docetaxel in immunogenic TNBC (FinHER) implies that Docetaxel interferes with lymphocytes in eliciting its anti-tumor effect. Supporting this idea, in the neoadjuvant setting, immunogenic TNBCs are less responsive to a taxane-containing regimen than regimens without MTAs (Figure 3A). Hence, the present study put forward the notion that taxanes may be limited to non-immunogenic tumors. Preclinical studies using whole tumor cell-based vaccines (tumor-specific granulocyte/macrophage-colony stimulating factor (GM-CSF)-secreting cells based vaccines) supported this notion by showing that microtubule-stabilizing agents were not effective in reducing tumor burden when administered after vaccination, while their pre-vaccine administration significantly decreased tumor burden. Specifically, in a neu-expressed trangenic breast cancer mouse model with GM-CSF-secreting and HER2/neu-expressing vaccine, pre-vaccine administration of Paclitaxel significantly reduced tumor burden compared to Paclitaxel only administration, whereas post-vaccine administration of Paclitaxel did not change tumor burden compared to Paclitaxel only regimen (44). The authors further demonstrated that, compared to the vaccination-only arm, pre-vaccine administration of Paclitaxel increased the neu-specific T-cell pool in the spleen, whereas post-vaccine administration of Paclitaxel significantly reduced it (44). Likewise, in the lung cancer mice models with GM-CSF-producing lung cancer vaccine, pre-vaccine administration of Docetaxel resulted in complete tumor regression in 70% of mice, whereas post-vaccine administration of Docetaxel did not reduce tumor burden in the mice tested (45). The authors further demonstrated that tumor regression is mediated by tumor-specific CD8 T-cells, not by CD4 T-cells, such that CD8 T-cell depletion abolishes tumor regression. The above study demonstrated that pre-vaccine administration of Docetaxel to non-immunogenic lung tumors (no tumor-specific CD8 T-cells before vaccination) augments the vaccine effect, whereas post-vaccine administration of Docetaxel to immunogenic lung tumors (presence of tumor-specific CD8 T-cells after vaccination) abolishes tumor regression. Collectively, the above preclinical studies demonstrated that microtubule-stabilizing agents lack efficacy in tumors with pre-existing immune responses induced by vaccines, but augment the vaccine effect when given before vaccination where there is no pre-existing tumor-specific immune response. Hence microtubule stabilizing agents would be an ideal chemo backbone for ICI in non-immunogenic TNBC.

Notably, significant heterogeneity in immune response measures between TIL density (TIL-H&E) and TIL-BP gene modules exists in the FinHER dataset. Specifically, none of the TIL-BP gene modules but the TIL-H&E counts showed a significant prognostic association in FinHER adjuvant TNBC (Supplementary Figure 15A) (3). Conversely, only the TIL-BP gene modules, but not the TIL-H&E counts, showed an interaction with MTAs in FinHER adjuvant TNBC (Figure 1A and 2, Supplementary Figures 7-13). In particular, TIL-H&E counts could not detect the potential taxane resistance associated with immunogenic TNBC that the TIL-BP gene modules detected (Figure 2, Supplementary Figures 9-13). This heterogeneity in results could be due to cellular phenotype dependence on environmental cues (46). TIL constitutes a heterogeneous collection of lymphocytes with their pro- or anti-cancer phenotypes determined by the tumor microenvironment including cancer cells (4,46,47). Hence a crude measure of TIL density cannot be interpreted as a functional anti-tumor response. Rather, it simply represents the degree of immunogenicity or the tumor’s dependency on an inflamed microenvironment for survival (48). Molecular signatures may be a better indicator of active signaling.

Intriguingly, contrary to interferon gene modules, immune gene modules’ interaction with MTAs was inconsistent in neoadjuvant and adjuvant TNBC; no significant interaction in neoadjuvant TNBC but significant interaction in adjuvant TNBC (Figures 1A and 3A, Supplementary Figure 14A). One potential reason is the difference between the adjuvant and neoadjuvant settings. The adjuvant setting focuses on eliminating dormant or active micro-metastasis after surgically removing the primary tumor. The high interferon and immune signaling in the surgically removed tumor represent the host’s ability to mount an immune response against the primary tumor. The immune response/TIL in these pretreatment resection specimens simply indicates that the tumor exploited the inflammatory microenvironment for its own survival. In other words, the cytotoxic potential of the immune response to these tumors is likely neutralized in one way or another before resection. It must be noted, however, that in the neoadjuvant setting, the primary tumor is present within the host, which can neutralize the cytotoxicity of the immune response against it (in specimens with high immune response/TIL). Therefore, interferon signaling, which has strong immunomodulatory capabilities (49,50), may have more clinical relevance than the compromised immune signaling present in unresected primary tumors in neoadjuvant settings.

The similarity of the direction of interaction between MTA and immune response on breast cancer patient survival in HR+BC and TNBC is intriguing, given the breast cancer subtype-specific differences in immune signaling and immune cell composition (51) and the differential effect of interferon signaling in TNBC and HR+BC (52,53). For instance, interferon treatment augments the immune response in ER negative tumors whereas in ER+ve tumors it suppresses the immune response by down-regulating HLA class II expression. However, contrary to the above report, increased interferon signaling is associated with a better response to non-MTA chemotherapy regimens in neoadjuvant HR+BC. Additionally, in contrast to neoadjuvant TNBC, the immune gene module significantly interacts with MTAs in neoadjuvant HR+BC, again highlighting subtype-specific immune signaling differences. The above observations call for additional research to carefully delineate the interaction between ER signaling, immune response, and MTAs in breast cancer.

### Limitations

In the present study, only TNBC is analyzed in both the test (FinHER adjuvant) and validation (pooled neoadjuvant GEO) datasets. HER2+BC is analyzed only in the adjuvant FinHER dataset and HR+BC only in the pooled neoadjuvant dataset.

No patients from destabilizing MTA regimens were present in the pooled neoadjuvant GEO dataset, possibly due to the omnipresence of taxanes in early breast cancer settings (35).

Dexamethasone, an immunosuppressive corticosteroid, was given to all patients receiving Docetaxel in the FinHER trial (16). Although dexamethasone is a lymphodepleting agent, it is reported to have no significant impact on the clinical outcome of neoadjuvant TNBC patients retrospectively (54). Corticosteroid administration is not considered when characterizing treatment regimens during GEO dataset integration.

Based on the consistency of probe level expression in FinHER expression profiles, a large number of probesets were discarded. This limits the application of insilico immune cell type estimation from FinHER due to the unreliability of cell-type marker gene modules in the FinHER dataset. As a result, the heterogeneity observed between TIL density and gene modules could not be investigated at the cellular level.

TIL counts are not available from the pooled neoadjuvant GEO dataset, but immune cell-type estimates were available. The T- and B-cell estimates from the above dataset are correlated better with immune gene modules than interferon gene modules (Supplementary Figure 6B), implying the immunomodulator role of interferon signaling (49,50). In line with this, the T-cell estimates displayed consistent results with immune gene modules, but not with interferon gene modules (Supplementary Figure 14) (49,50).

The GEO dataset is not randomized and integrates molecular data from studies that used multiple drugs of the same drug classes, different patient selection strategies, pCR definitions, sample procurement methods, and gene expression profiling platforms to increase the sample size. These dataset-specific effects, however, were accounted for by a dataset-id term in logistic regression models.

Chemotherapy’s impact on the immune system varies among drugs and is evident in drug-dependent immune cell expansion or depletion (55). In the FinHER dataset the impact of FEC on the immune cell population in FinHER TNBC is unclear. However, the differential effect observed can be directly attributed to Docetaxel/Vinorelbine due to the randomized setting.

Further, in the neoadjuvant GEO dataset, the analysis is limited to Anthracycline, Antimetabolite, and Alkylating agent containing regimens. This is to control bias introduced by polytherapy regimens with various treatment combinations, and also to make the treatment regimen congruent with the FinHER dataset’s FEC regimen. Besides the drug-specific effects on immune cells mentioned previously, the sequence and dosage of drug administration have profound effects on modulating immune responses (56). These factors are not considered in the analysis. Despite pooling multiple drugs of the same class in the GEO dataset, the direction of interaction between stabilizing agents versus no microtubule agent was in line with FinHER results, providing increased confidence.

Finally, the present retrospective study used bulk tumor expression data. As a result, immune gene modules and TIL density only indirectly suggest the presence of multiple T cell subtypes. Therefore, the effects of microtubule-stabilizing/destabilizing agents on T-cells are speculative. A preclinical study using immunogenic and non-immunogenic TNBC model systems with microtubule-stabilizing/destabilizing agents and simultaneous transcriptome and proteome profiling at the single-cell level (57) is essential to validate the results and gain more direct insights into microtubule biology on immune cells, particularly at the post-transcriptional level.

## Conclusion

Among the breast cancer data analyzed, TNBC and HER2+BC from the adjuvant FinHER dataset and TNBC and HR+BC from the neoadjuvant dataset integrated from GEO, MTAs influence the clinical benefit associated with immune response measured by gene modules in the adjuvant/neoadjuvant TNBC and neoadjuvant HR+BC, but not in adjuvant HER2+BC. Heterogeneity exists in host immune response measurements using TIL-H&E and gene modules. The marked heterogeneity could be due to the dependence of cellular phenotype on its surrounding environment, which may confound the host immune response measured by cell-type enumeration methods. The observation that Docetaxel, a stabilizing MTA, gives a worse response and Vinorelbine, a destabilizing MTA, gives a better response in immunogenic TNBC has relevance in disease management and clinical trial design. Moreover, the present study provides evidence for bringing Vinorelbine, currently limited to the metastatic breast cancer setting, to the early immunogenic TNBC setting. However, the limited size of datasets used to fit interaction models warrants additional analyses.

## Author contributions

Concept and design of the work: Vinu Jose and Christos Sotiriou

Data acquisition: Francoise Rothe, Debora Fumagalli, Samira Majjaj, Delphine Vincent, Roberto Salgado, Nicolas Sirtaine, Sherene Loi, Heikki Joensuu, and Christos Sotiriou Data curation: Vinu Jose

Data analysis: Vinu Jose, David Venet, and Stefan Michiels Data interpretation: All authors

Resources: Heikki Joensuu and Christos Sotiriou Funding acquisition and supervision: Christos Sotiriou Original draft preparation: Vinu Jose

Manuscript review and editing: All authors

## Data Availability

The clinical data of the FinHER trial are protected by Finnish law. To use FinHER clinical data for research, the intended study plan must be approved by the ethics committee of Helsinki University Hospital (www.hus.fi), and comply with patient informed consent. The raw and reprocessed FinHER gene expression data can be accessed from GEO accession GSE47994. The integrated neoadjuvant GEO gene expression dataset can be accessed under GEO accession number GSE205568.

https://doi.org/10.17605/OSF.IO/URT9X

https://www.ncbi.nlm.nih.gov/geo/query/acc.cgi?acc=GSE47994

https://www.ncbi.nlm.nih.gov/geo/query/acc.cgi?acc=GSE205568

## Acknowledgements

The gene module validation analysis in full is based upon data generated by the TCGA Research Network: https://www.cancer.gov/tcga and Metabric consortium: https://www.cbioportal.org/study/summary?id=brca_metabric. The authors acknowledge the contribution of Debora Fumagalli during the data collection process. Vinu Jose received funding from the F.R.S-FNRS grant “Fonds de la Recherche Scientifique” – Télévie (Belgium), for this work (Credit no: 7461813F). The funders had no role in the study design, data collection, analysis, publication decision, or manuscript preparation. Roberto Salgado is supported by the Breast Cancer Research Foundation (BCRF, grant nr. 17-194). Sherene Loi is supported by the National Breast Cancer Foundation of Australia Endowed Chair and the Breast Cancer Research Foundation, New York. Heikki Joensuu is supported by the Sigrid Juselius Foundation, Finland.

Large multi-tab Excel file available separately.

Supplementary Table 1: TILsig. Table consists of (1) TILsig derived from TNBC, HER2+BC, and pooled TNBC and HER2+BC, (2) a list of over-represented BP terms, and (3) TIL-BP gene modules.

Large multi-tab Excel file available separately.

Supplementary Table 2: Published gene modules. Table consists of original gene modules, gene module subsets, gene module reliability test statistics, and gene module extraction details.

---

**Supplementary Figure 1:**
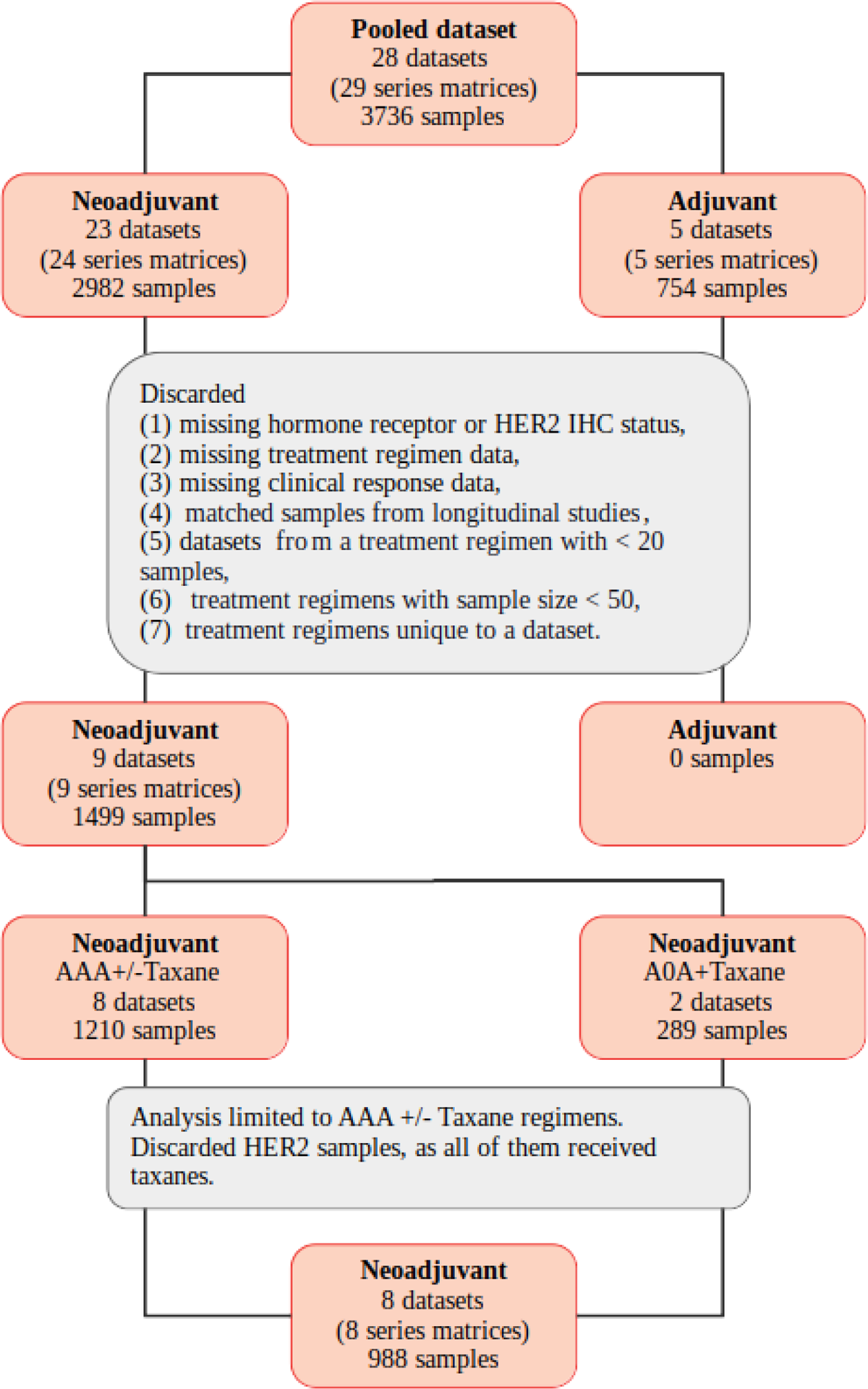
Additional sample/dataset filtering criteria for the GEO integrated dataset extracted from GSE205568.

**Supplementary Figure 2:**
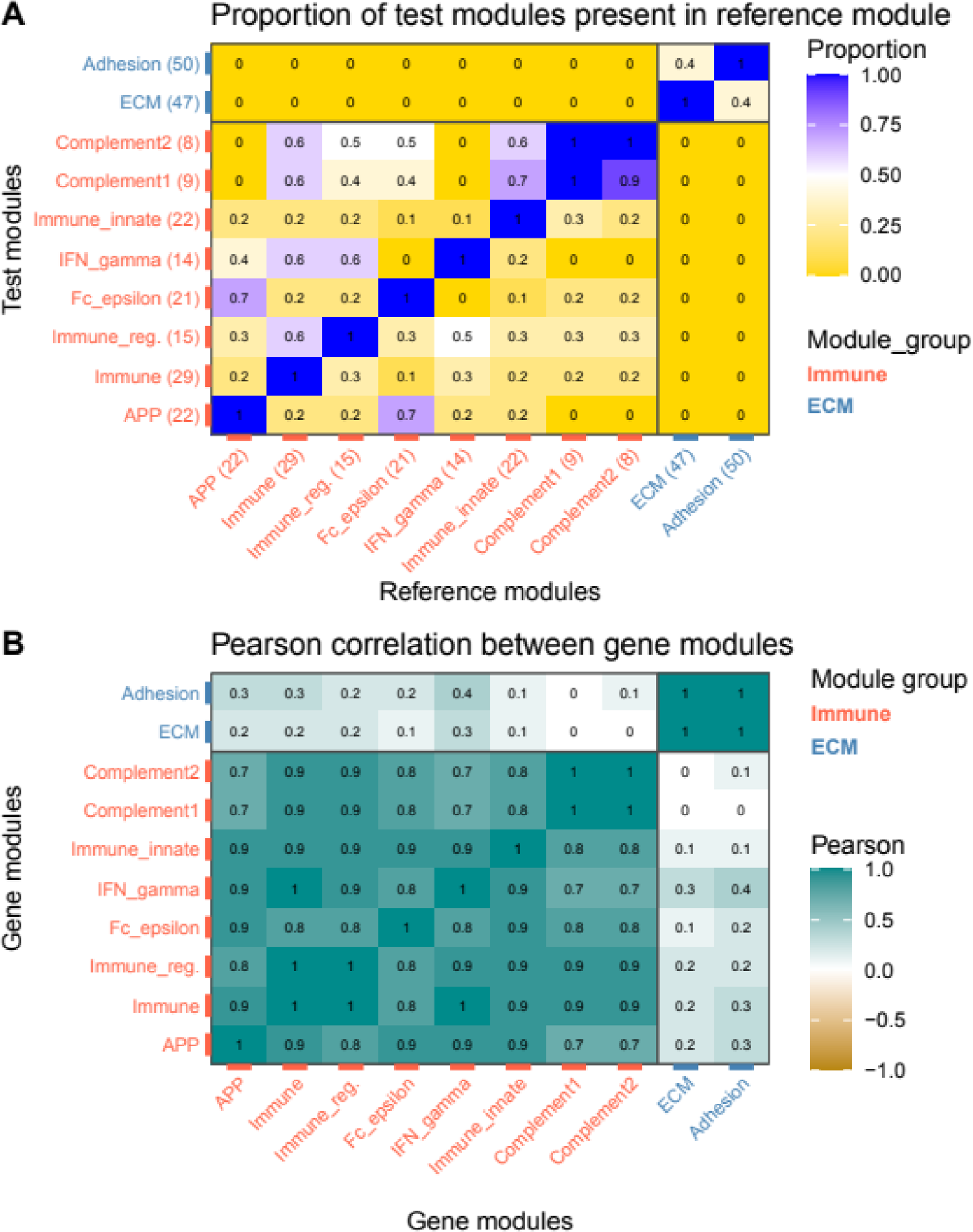
Gene overlap and module score correlations of de novo TIL associated biological-process gene modules. (A) Gene-module gene overlaps. Reference and test module designation is arbitrary and used for proportion calculation. The number within parenthesis next to the module name indicates the module size; (B) Module score correlation from the FinHER dataset.

**Supplementary Figure 3:**
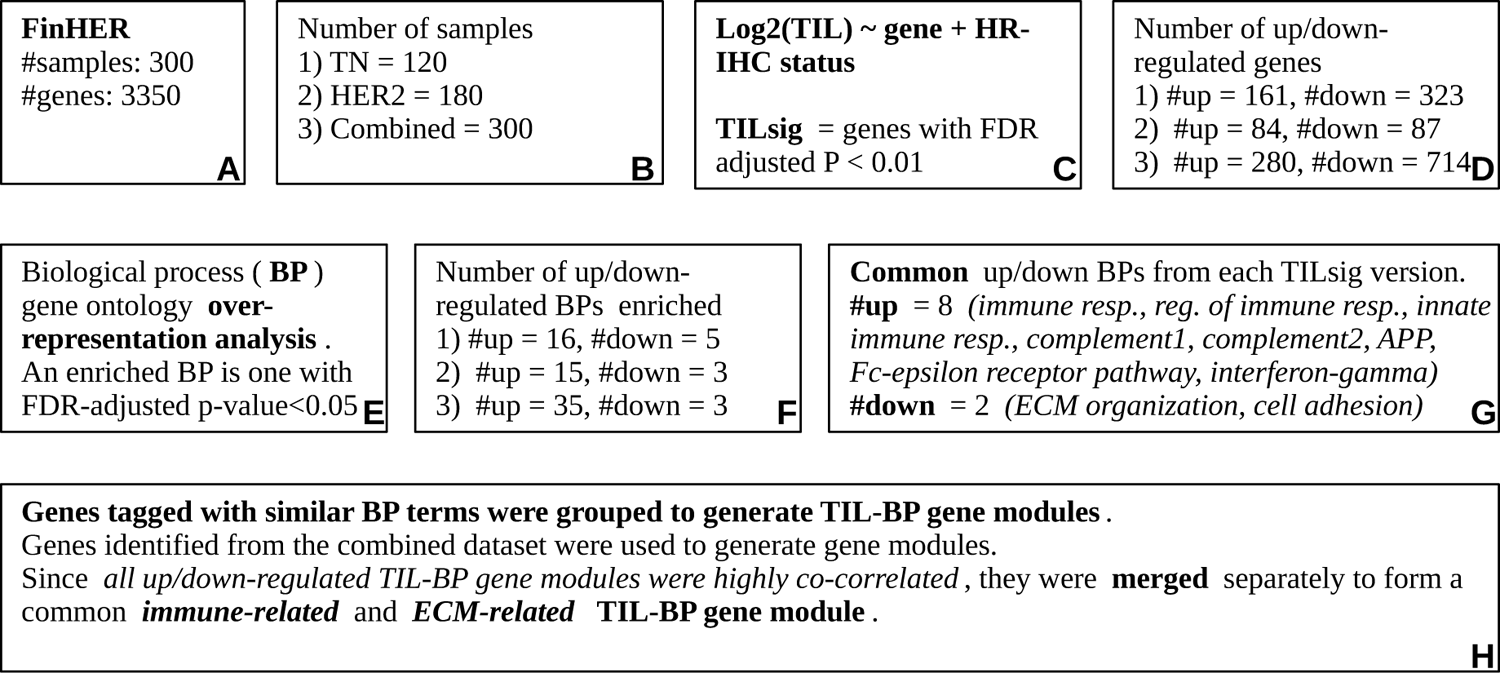
De novo TILsig generation workflow.

**Supplementary Figure 4:**
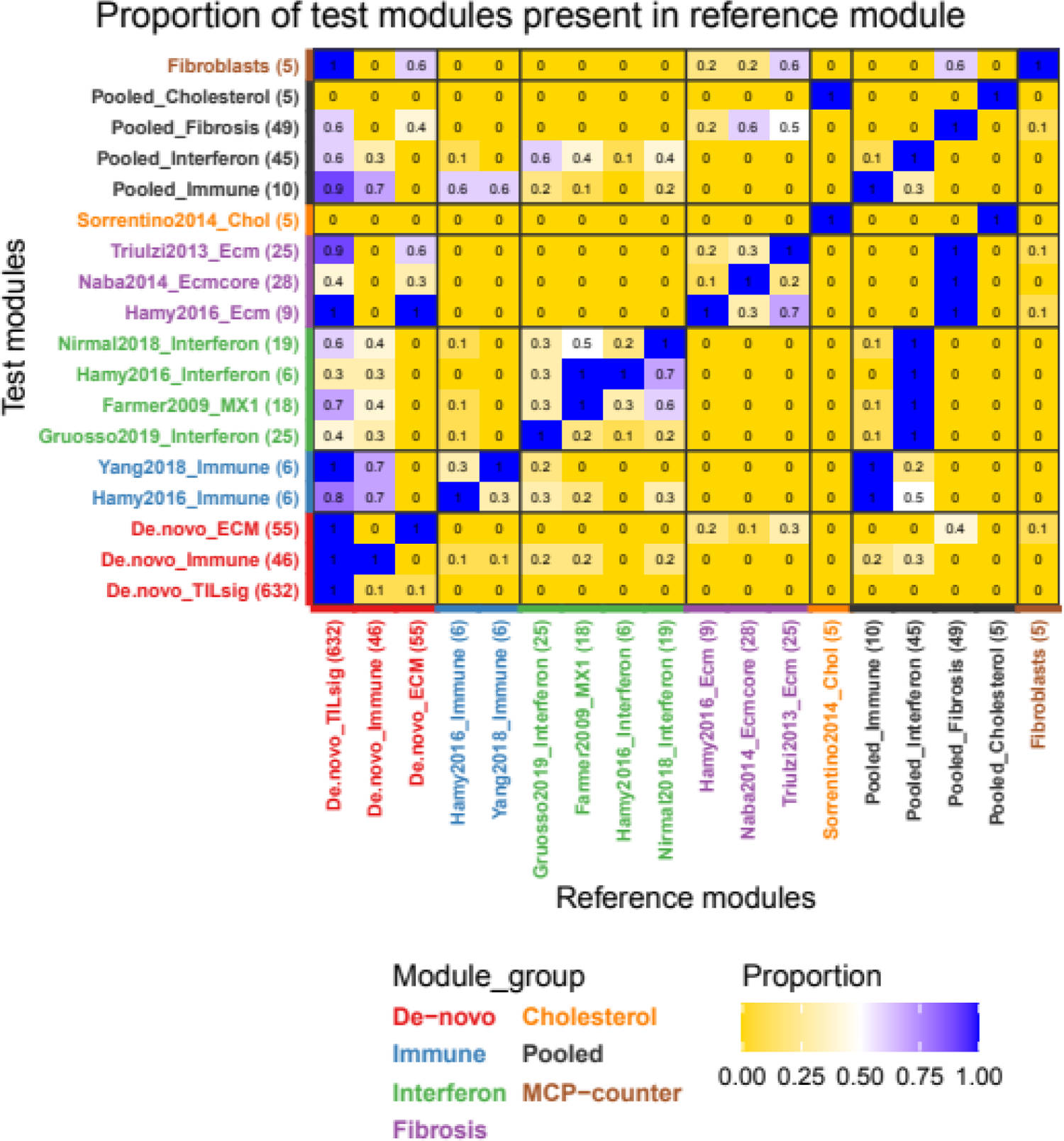
Gene overlap of FinGEO module subsets derived from published TIL-associated biological-process gene modules. Reference and test module designation is arbitrary and used for proportion calculation. The number within parenthesis next to the module name indicates the module size.

**Supplementary Figure 5:**
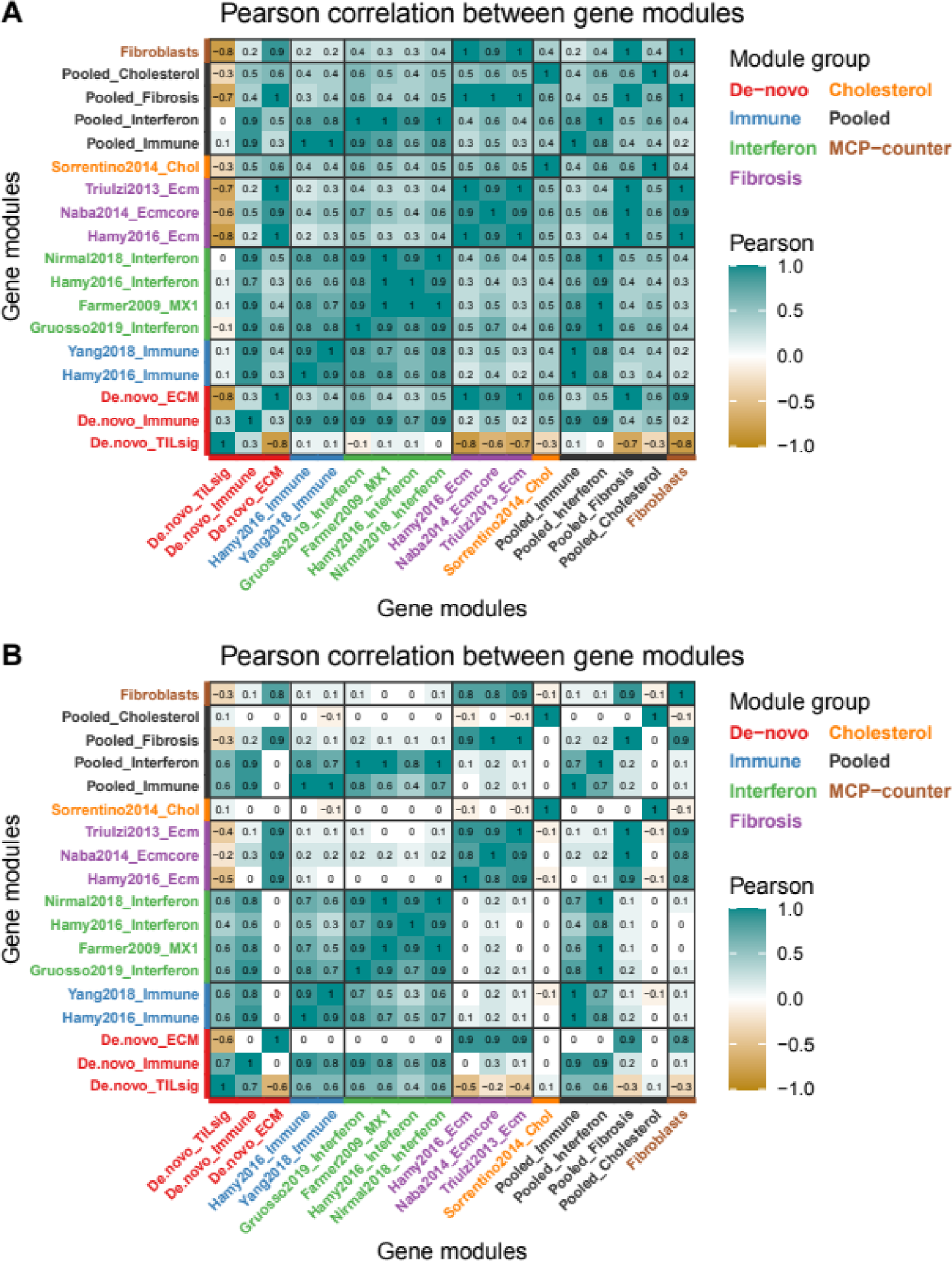
Module score correlations of FinGEO module subsets derived from published TIL-associated biological-process gene modules. Module score correlations from (A) FinHER and (B) integrated GEO datasets.

**Supplementary Figure 6:**
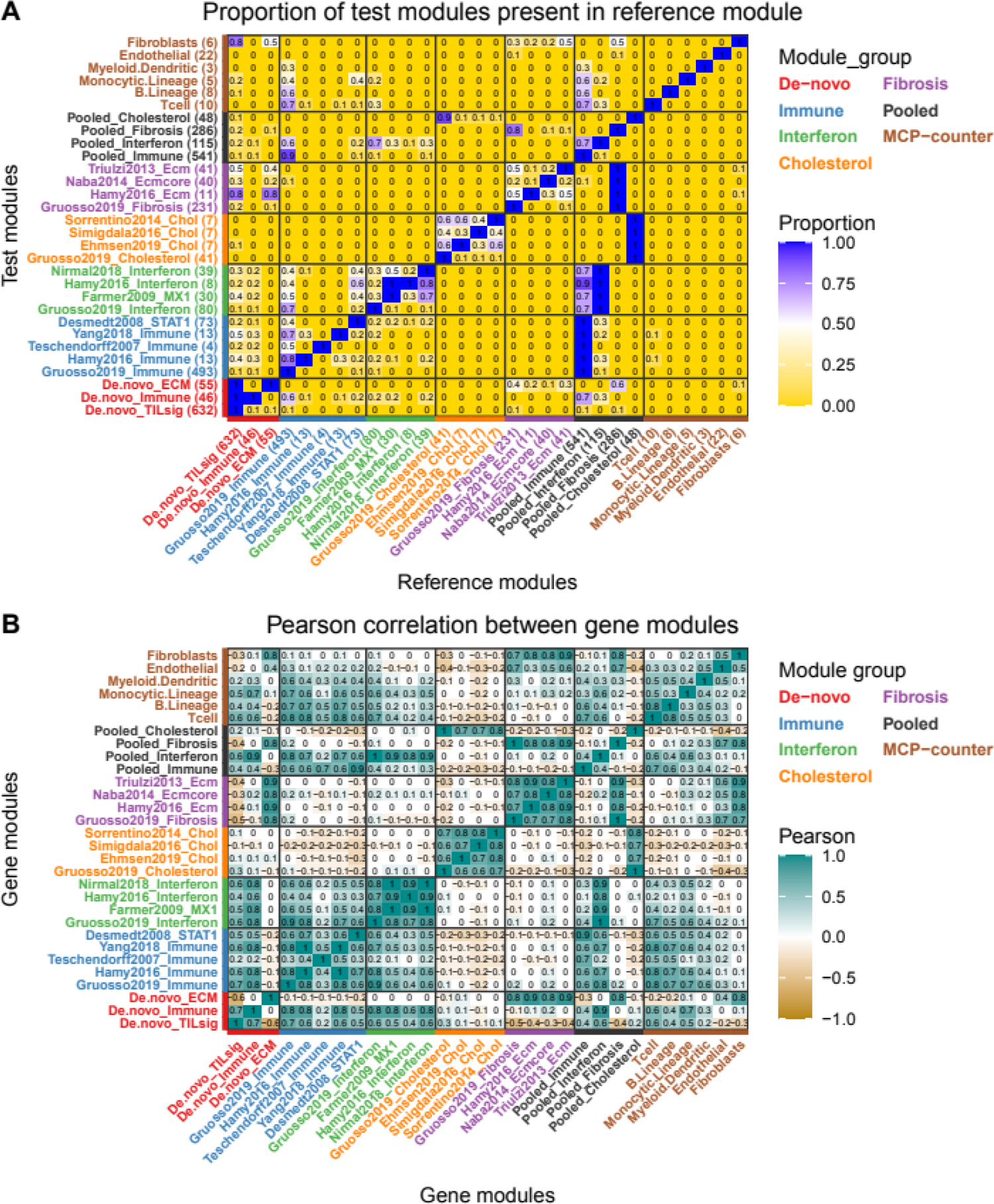
Gene overlap and module score correlations of GEO module subsets derived from published TIL-associated biological-process gene modules. (A) Gene module gene overlaps. Reference and test module designation is arbitrary and used for proportion calculation. The number within parenthesis next to the module name indicates the module size; (B) Module score correlation from the integrated GEO dataset.

**Supplementary Figure 7:**
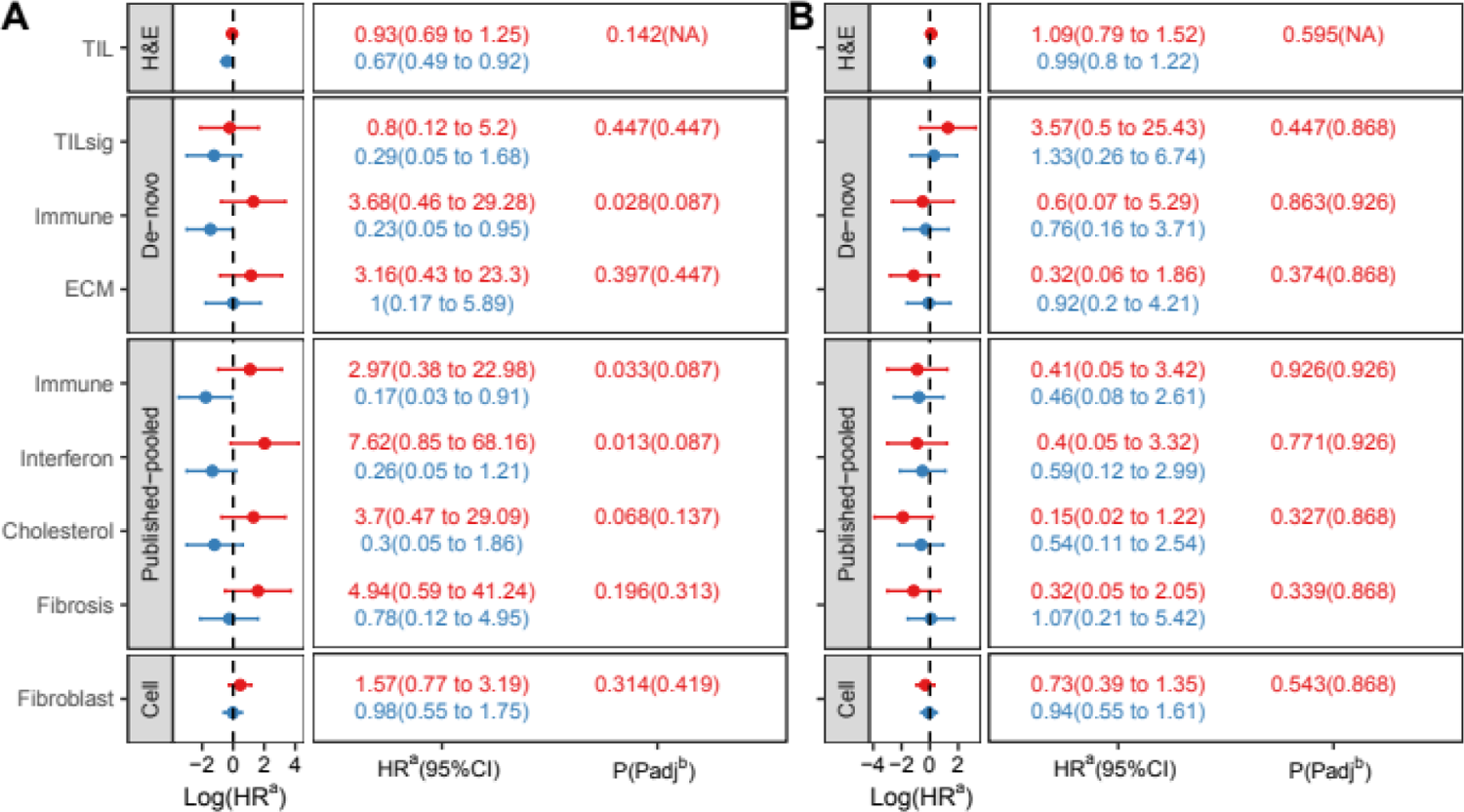
DDFS interaction between MTAs and immune response measures (TIL-H&E/FinGEO module subsets) in the FinHER adjuvant dataset. (A) TNBC; (B) HER2+BC. A positive log(HR) (i.e. HR>1) indicates a higher distant disease recurrence rate, while a negative log(HR) (i.e. HR<1) indicates a lower distant disease recurrence rate with an increase in module score in respective treatment regimens. Red represents Docetaxel - stabilizing MTA, and blue represents Vinorelbine - destabilizing MTA. ^a^ Hazard ratio. ^b^ TIL-H&E interaction p-value is not used for FDR correction.

**Supplementary Figure 8:**
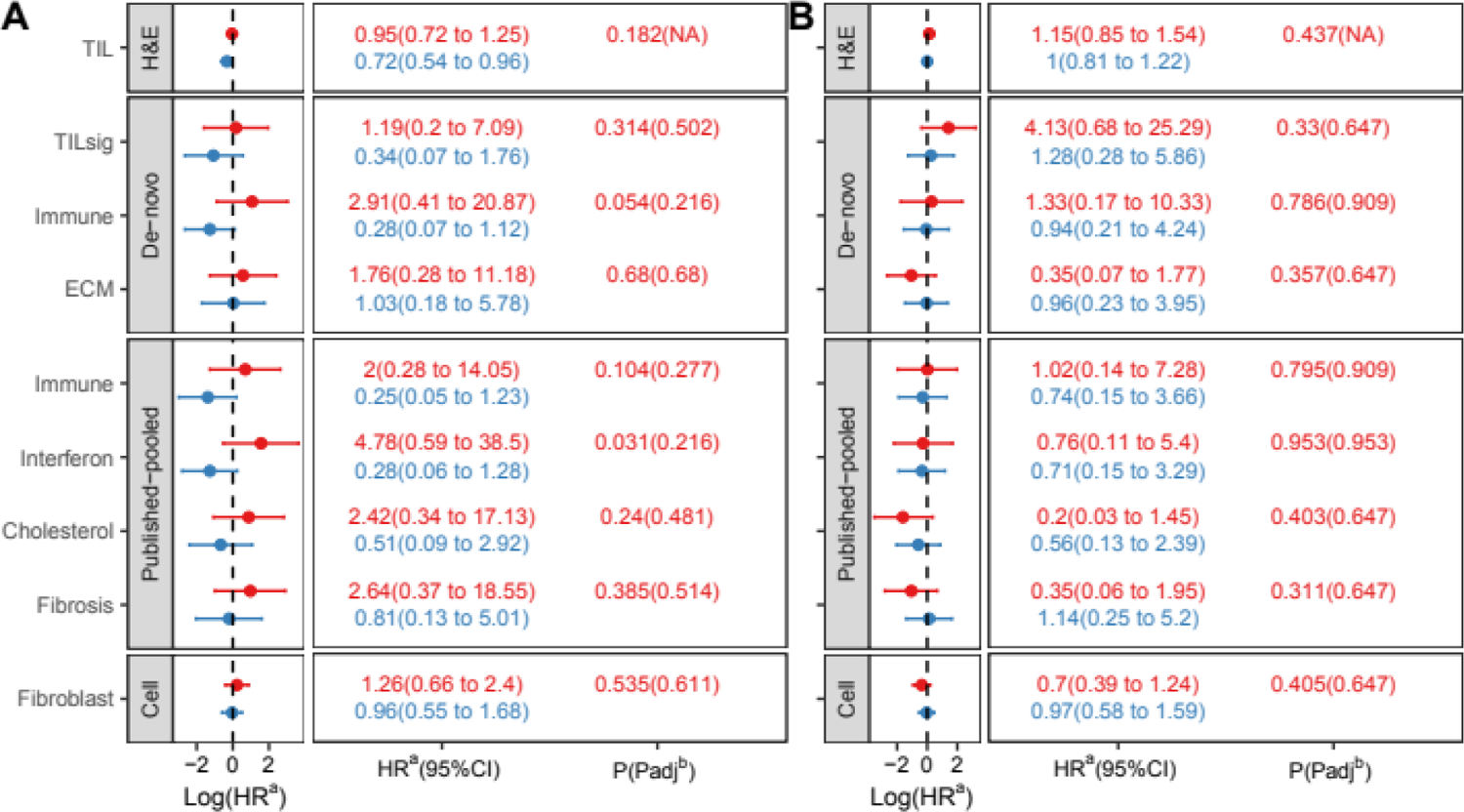
RFS interaction between MTAs and immune response measures (TIL-H&E/FinGEO module subsets) in the FinHER adjuvant dataset. (A) TNBC; (B) HER2+BC. A positive log(HR) (i.e. HR>1) indicates a higher disease recurrence rate, while a negative log(HR) (i.e. HR<1) indicates a lower disease recurrence rate with an increase in module score in respective treatment regimens. Red represents Docetaxel - stabilizing MTA, and blue represents Vinorelbine - destabilizing MTA. ^a^ Hazard ratio. ^b^ TIL-H&E interaction p-value is not used for FDR correction.

**Supplementary Figure 9:**
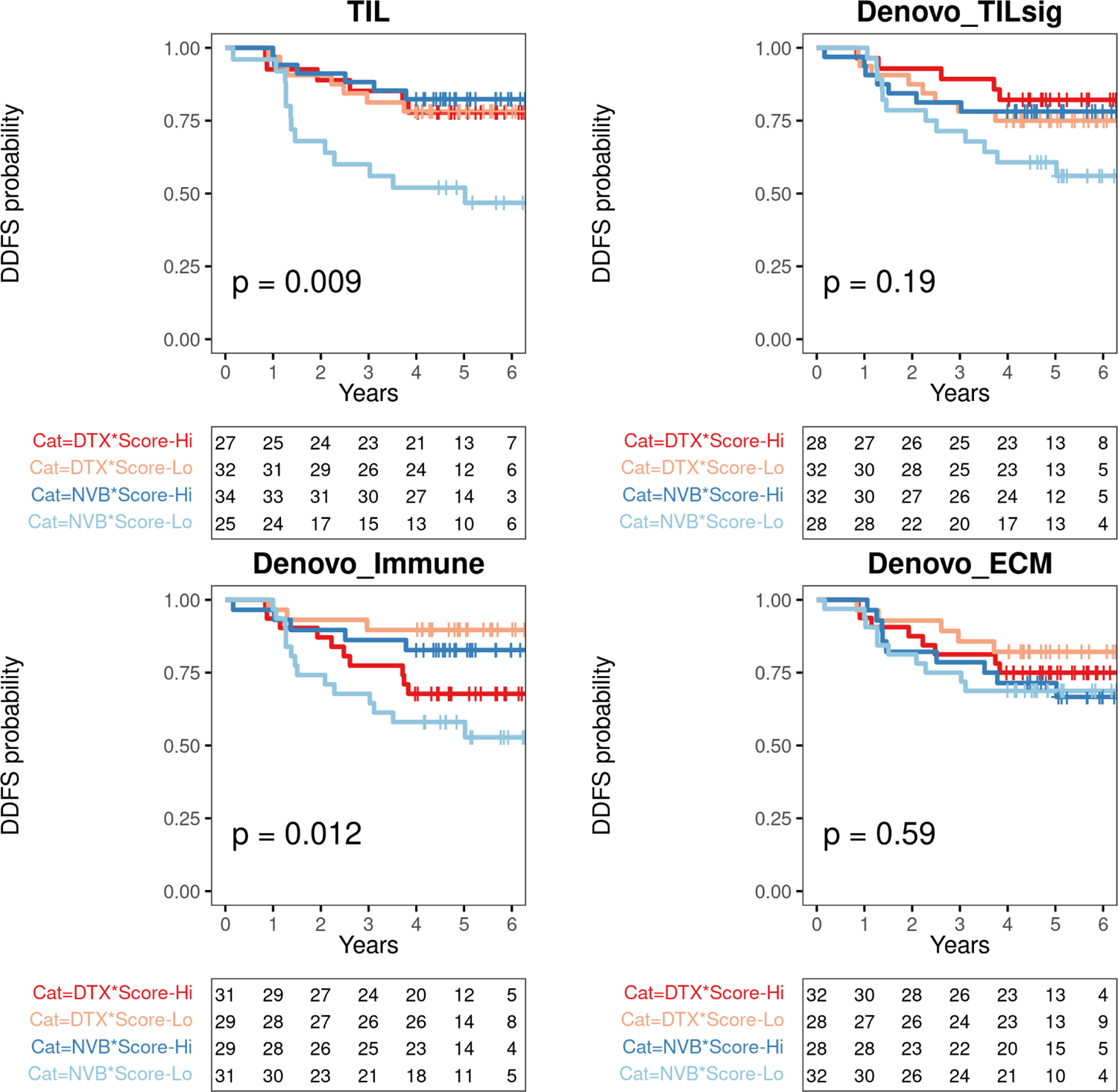
DDFS Kaplan-Meyer plots reveal the interaction between MTAs and selected immune response measures (TIL-H&E and de novo gene modules) in adjuvant FinHER TNBC. Dark and light red represent Docetaxel with high and low scores respectively. Dark and light blue represent Vinorelbine with high and low scores respectively. High/low classification is based on corresponding medians from TNBC. The median score for TIL (TIL-H&E) in TNBC is 25%. The P-value is computed using the Log-rank test. The table below each Kaplan-Meyer plot shows the number of patients at risk at each time point for each stratum. DTX: Docetaxel - stabilizing MTA. NVB: Vinorelbine - destabilizing MTA.

**Supplementary Figure 10:**
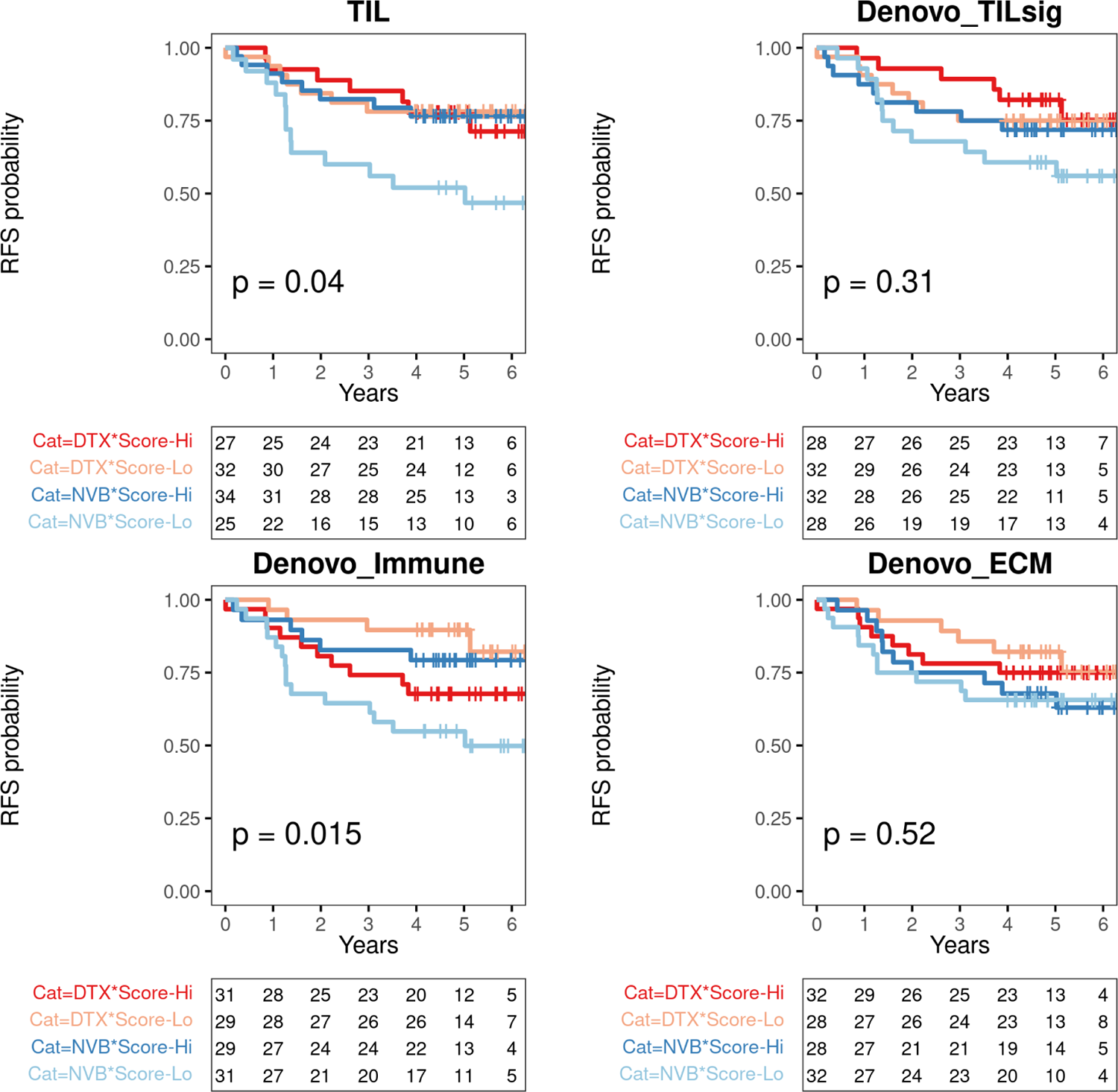
RFS Kaplan-Meyer plots reveal the interaction between MTAs and selected immune response measures (TIL-H&E and de novo gene modules) in adjuvant FinHER TNBC. Dark and light red represent Docetaxel with high and low scores respectively. Dark and light blue represent Vinorelbine with high and low scores respectively. High/low classification is based on corresponding medians from TNBC. The median score for TIL (TIL-H&E) in TNBC is 25%. The P-value is computed using the Log-rank test. The table below each Kaplan-Meyer plot shows the number of patients at risk at each time point for each stratum. DTX: Docetaxel - stabilizing MTA. NVB: Vinorelbine - destabilizing MTA.

**Supplementary Figure 11:**
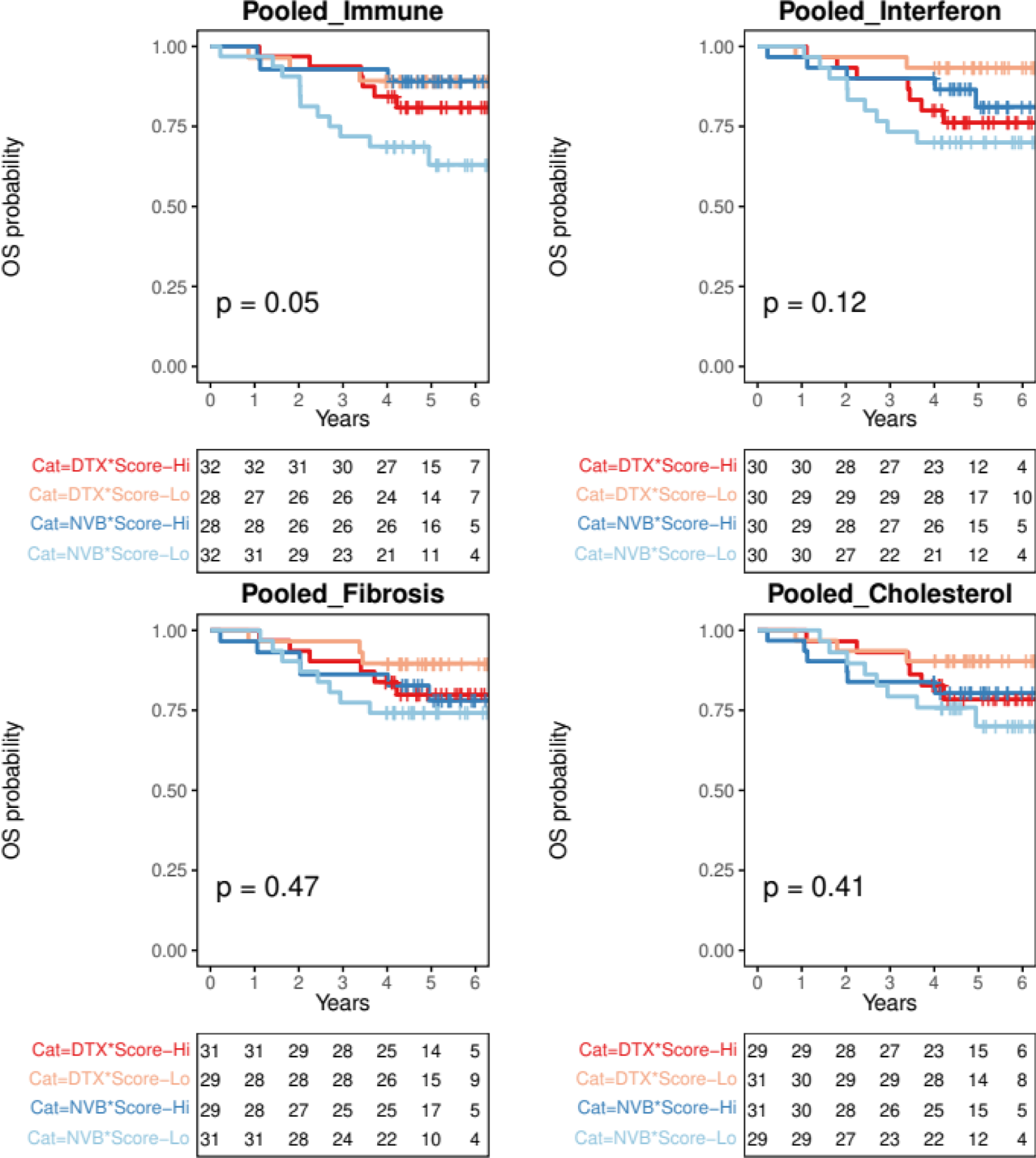
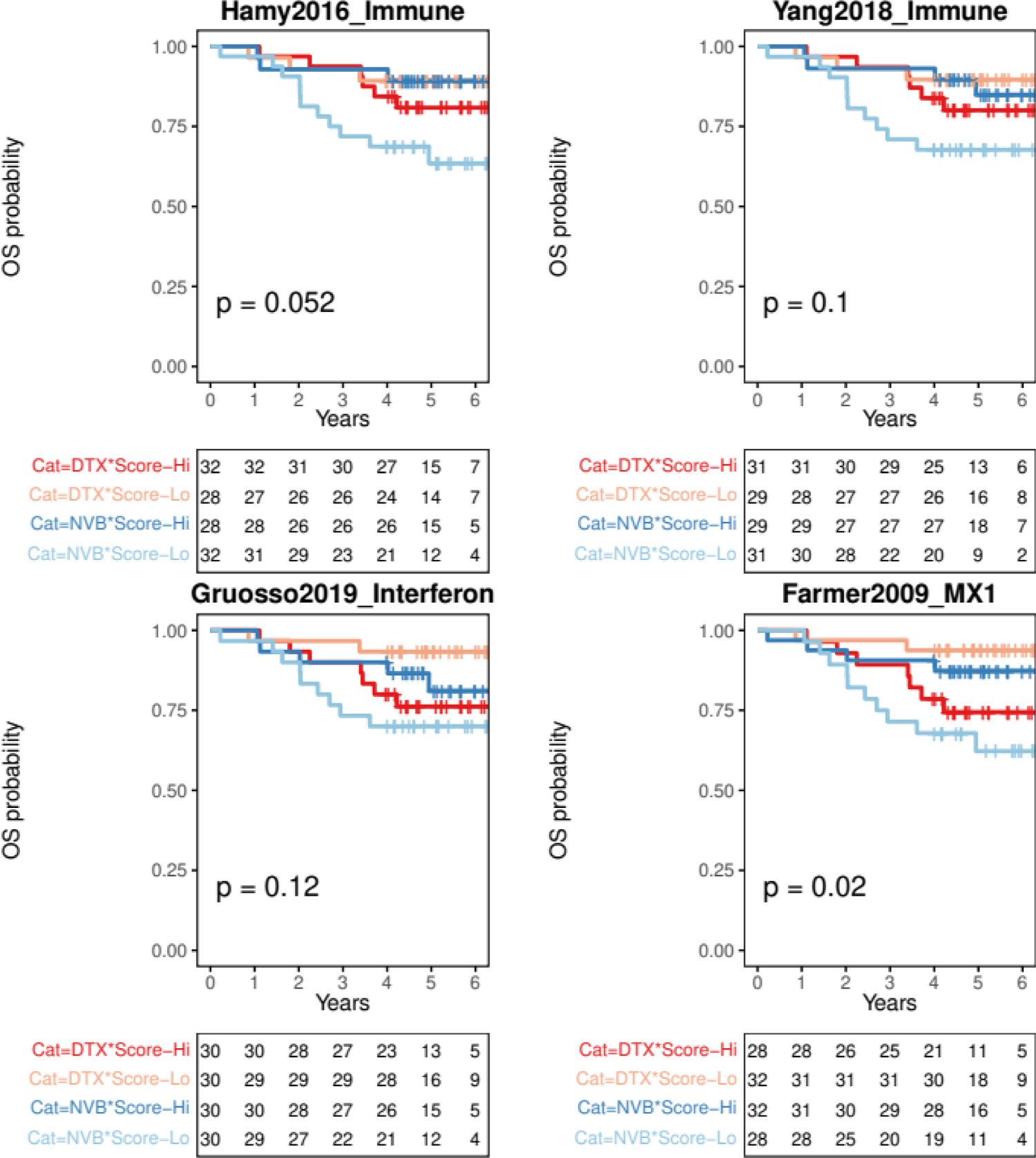
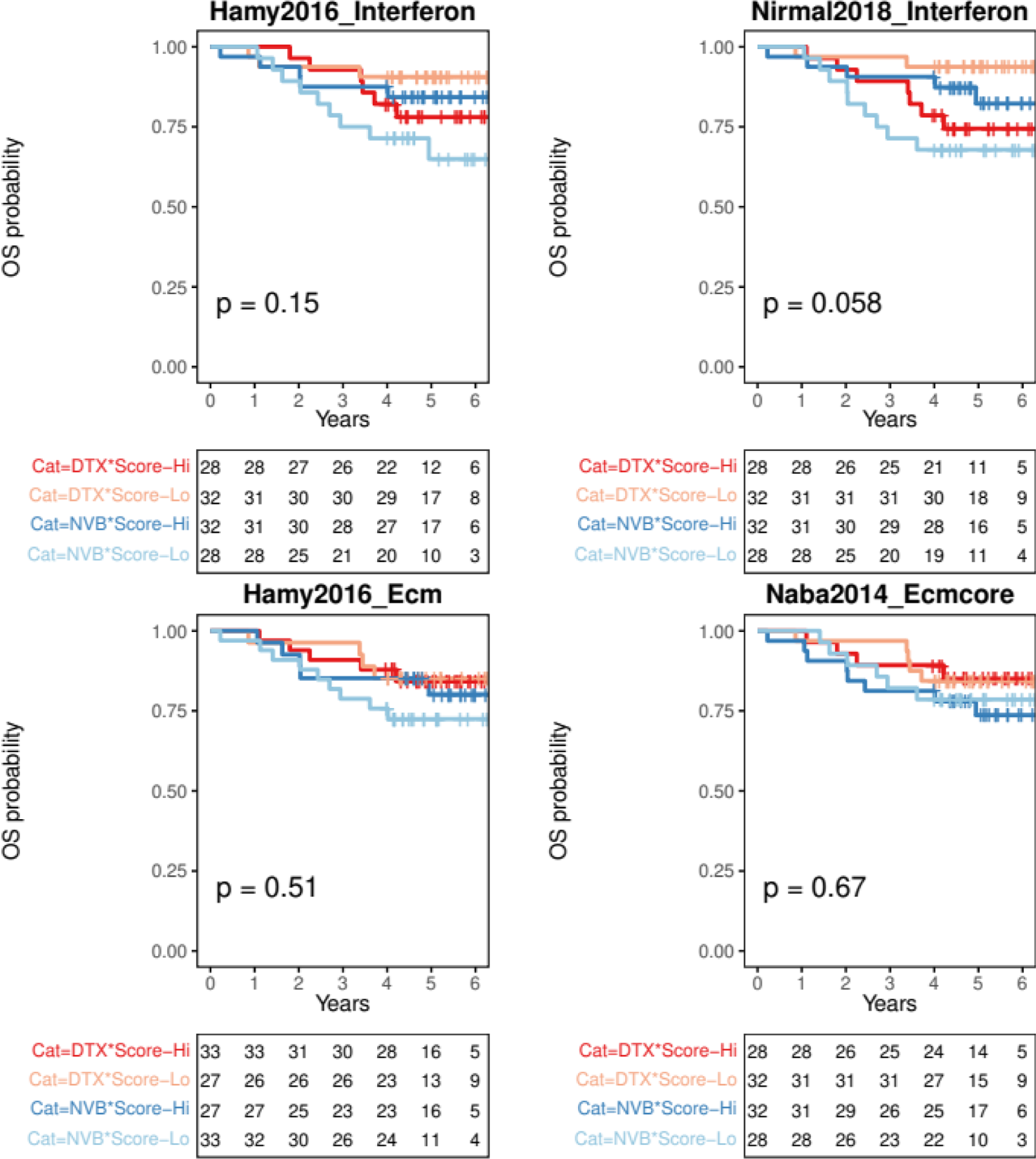
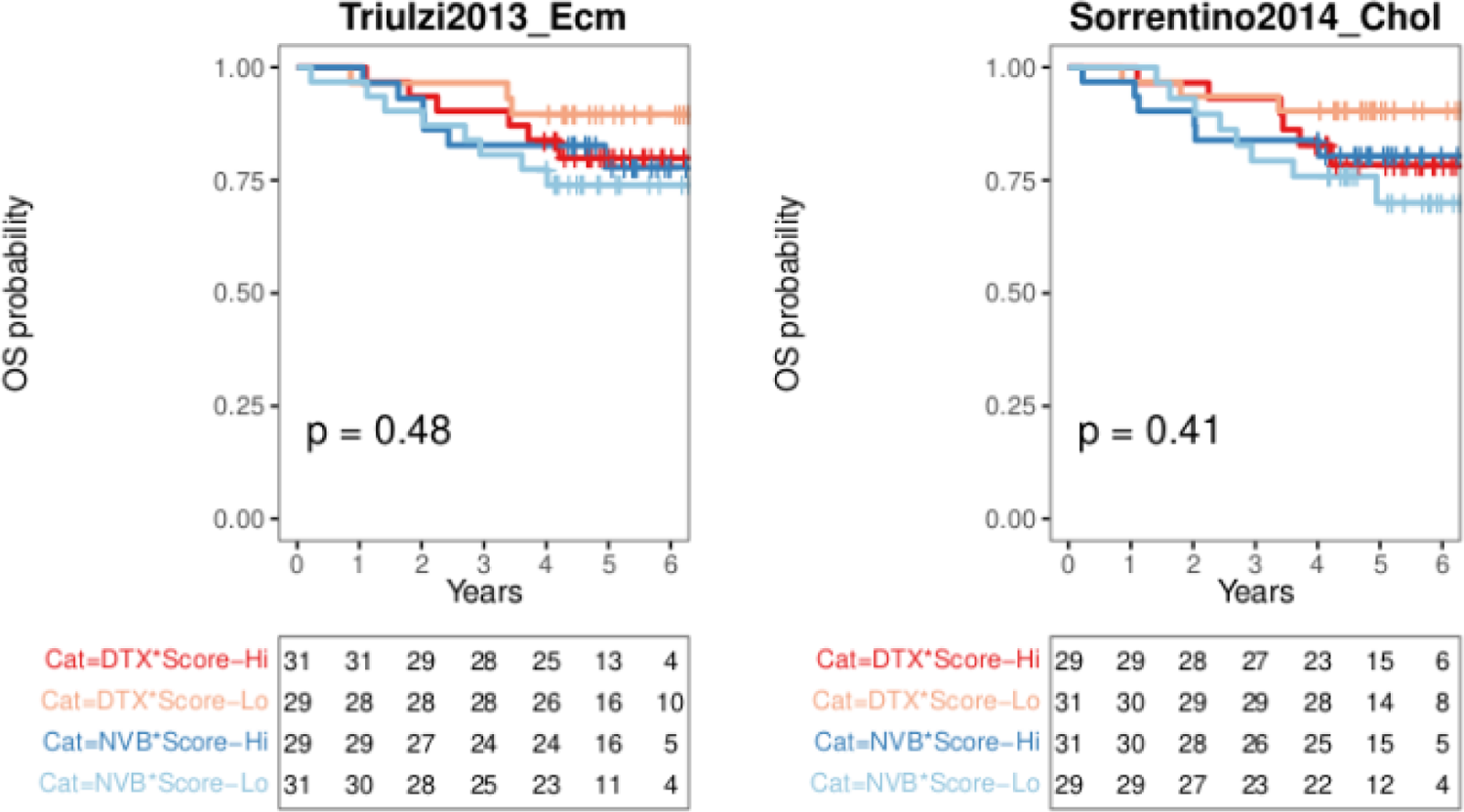
OS Kaplan-Meyer plots reveal the interaction between MTAs and immune response measures in adjuvant FinHER TNBC. Dark and light red represent Docetaxel with high and low scores respectively. Dark and light blue represent Vinorelbine with high and low scores respectively. High/low classification is based on corresponding medians from TNBC. The median score for TIL (TIL-H&E) in TNBC is 25%. The P-value is computed using the Log-rank test. The table below each Kaplan-Meyer plot shows the number of patients at risk at each time point for each stratum. DTX: Docetaxel - stabilizing MTA. NVB: Vinorelbine - destabilizing MTA.

**Supplementary Figure 12:**
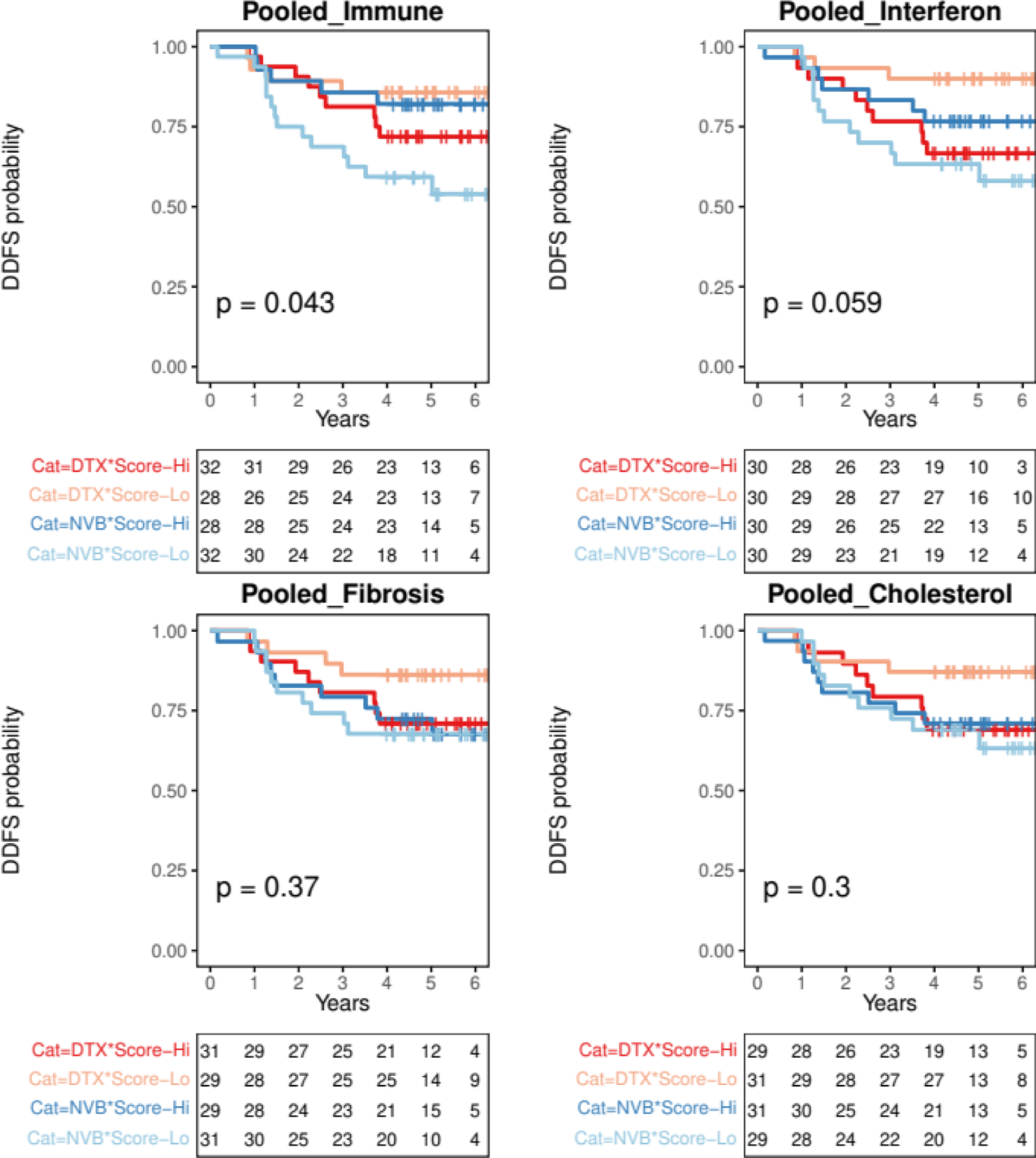
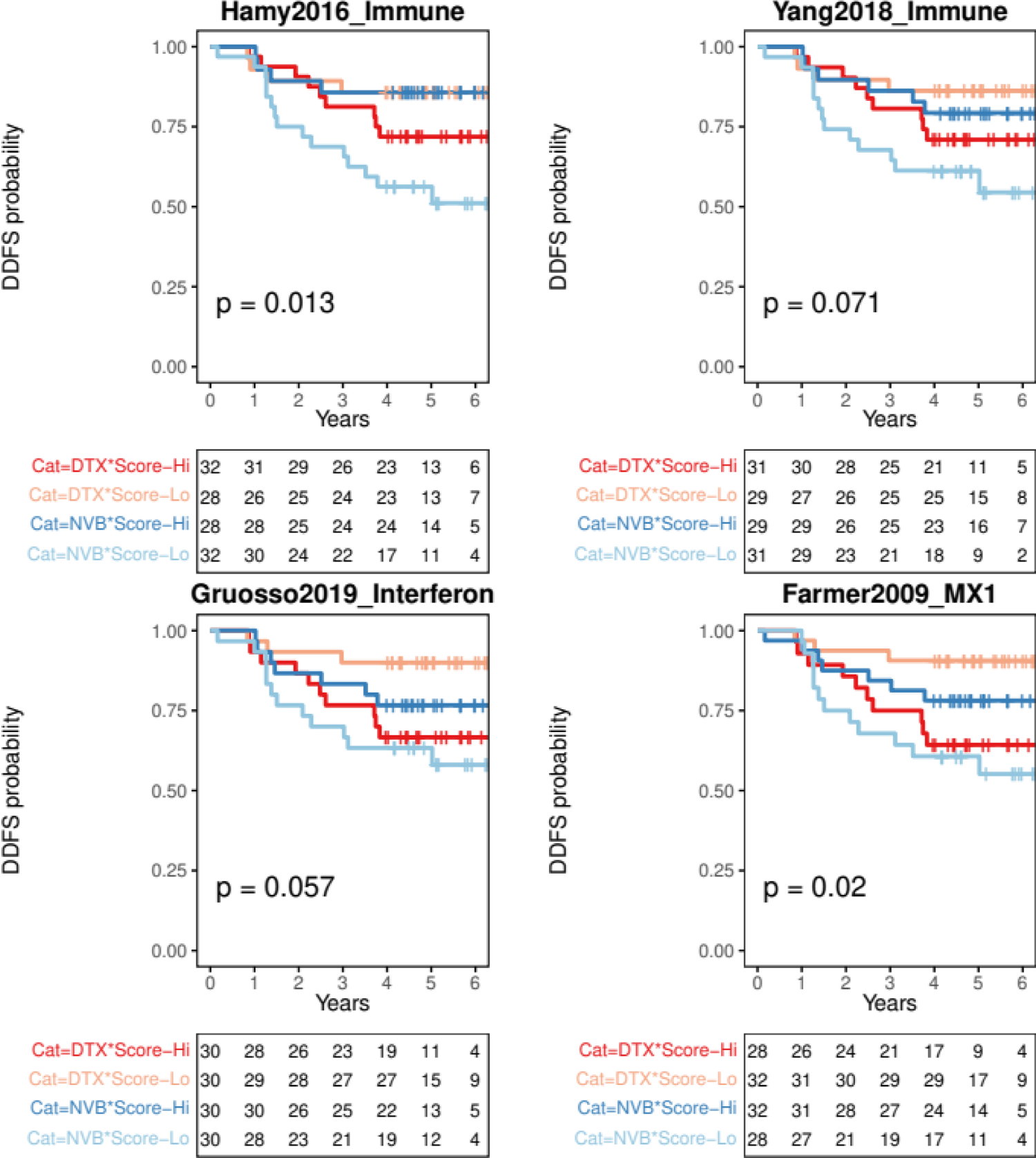
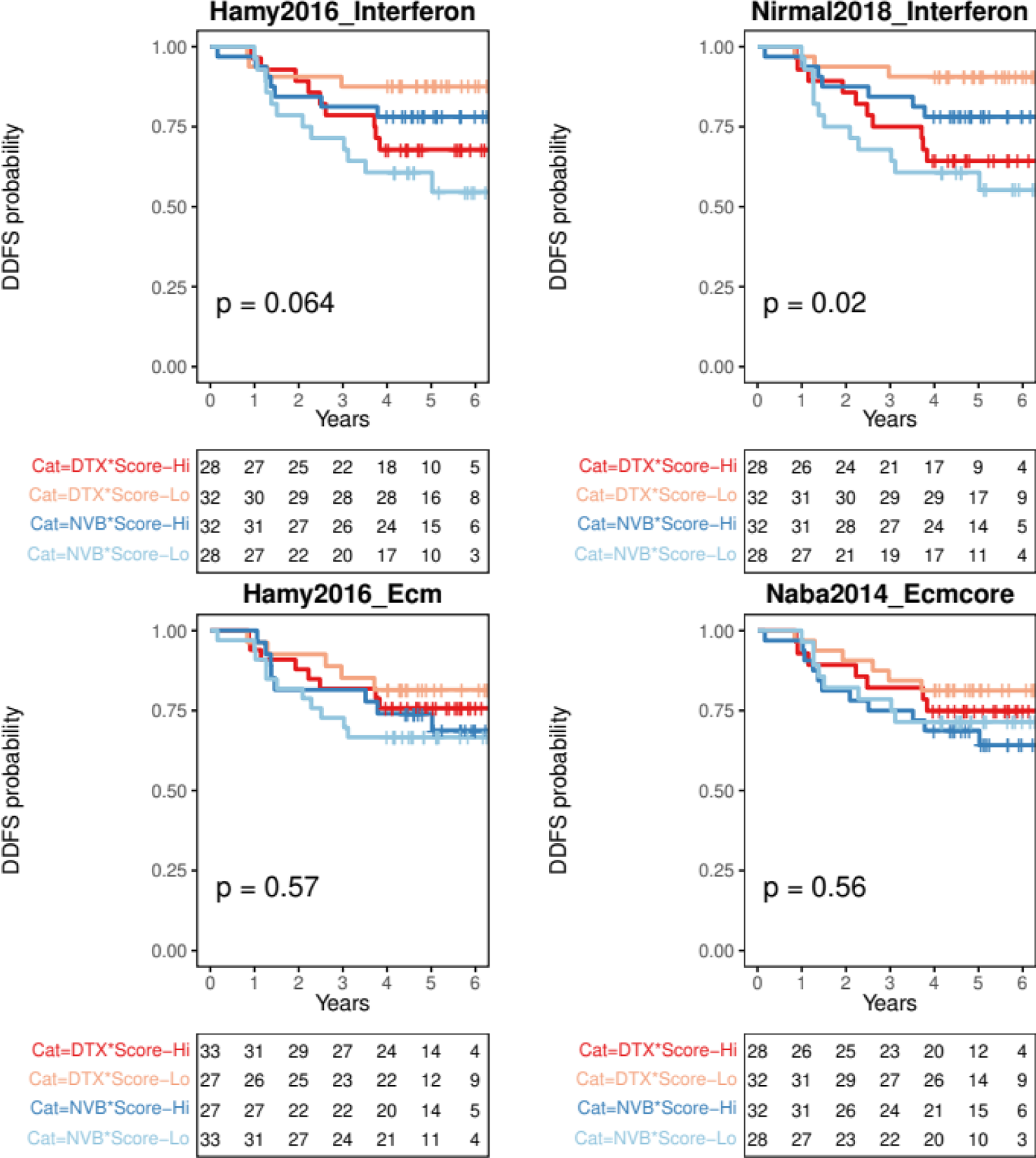
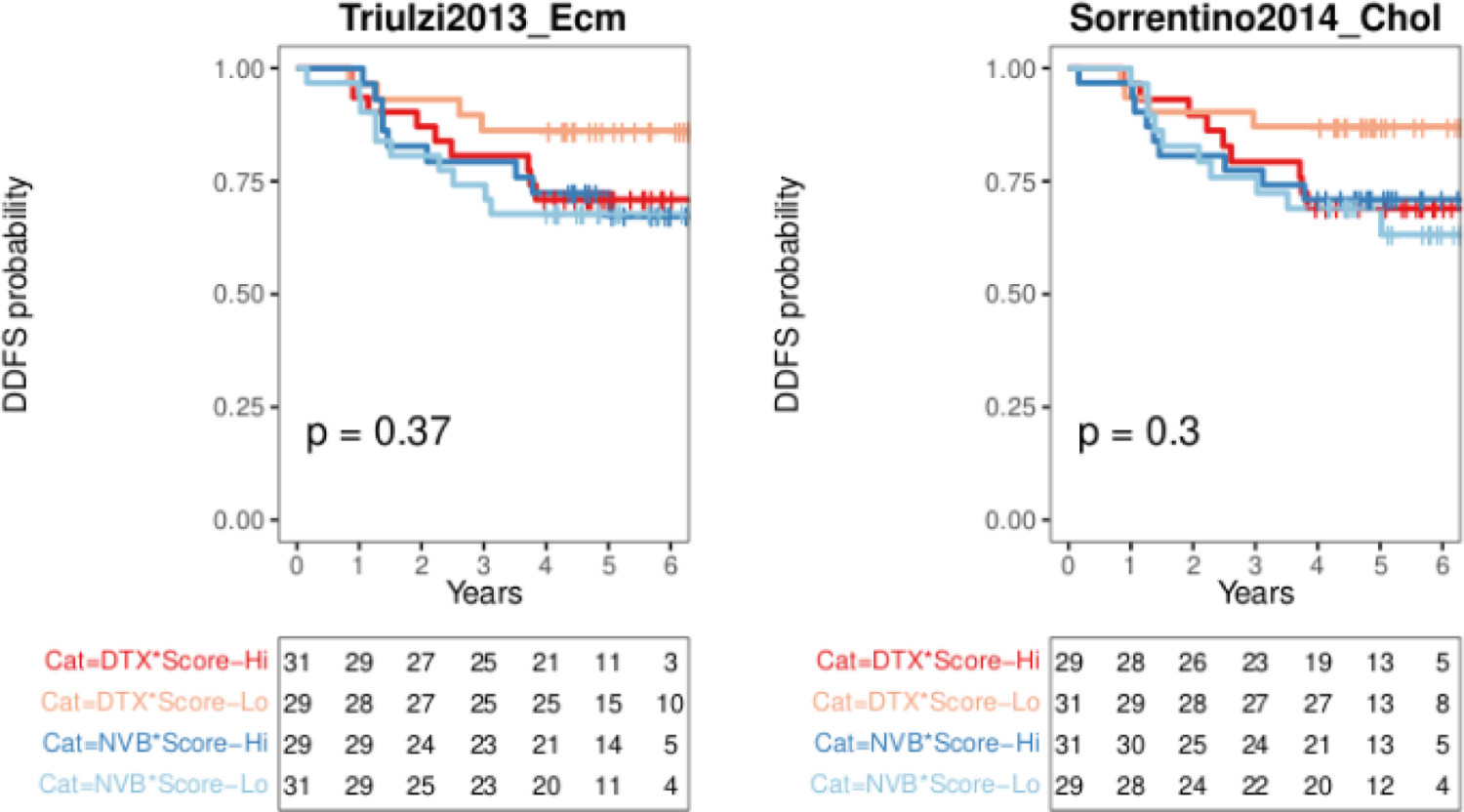
DDFS Kaplan-Meyer plots reveal the interaction between MTAs and immune response measures in adjuvant FinHER TNBC. Dark and light red represent Docetaxel with high and low scores respectively. Dark and light blue represent Vinorelbine with high and low scores respectively. High/low classification is based on corresponding medians from TNBC. The median score for TIL (TIL-H&E) in TNBC is 25%. The P-value is computed using the Log-rank test. The table below each Kaplan-Meyer plot shows the number of patients at risk at each time point for each stratum. DTX: Docetaxel - stabilizing MTA. NVB: Vinorelbine - destabilizing MTA.

**Supplementary Figure 13:**
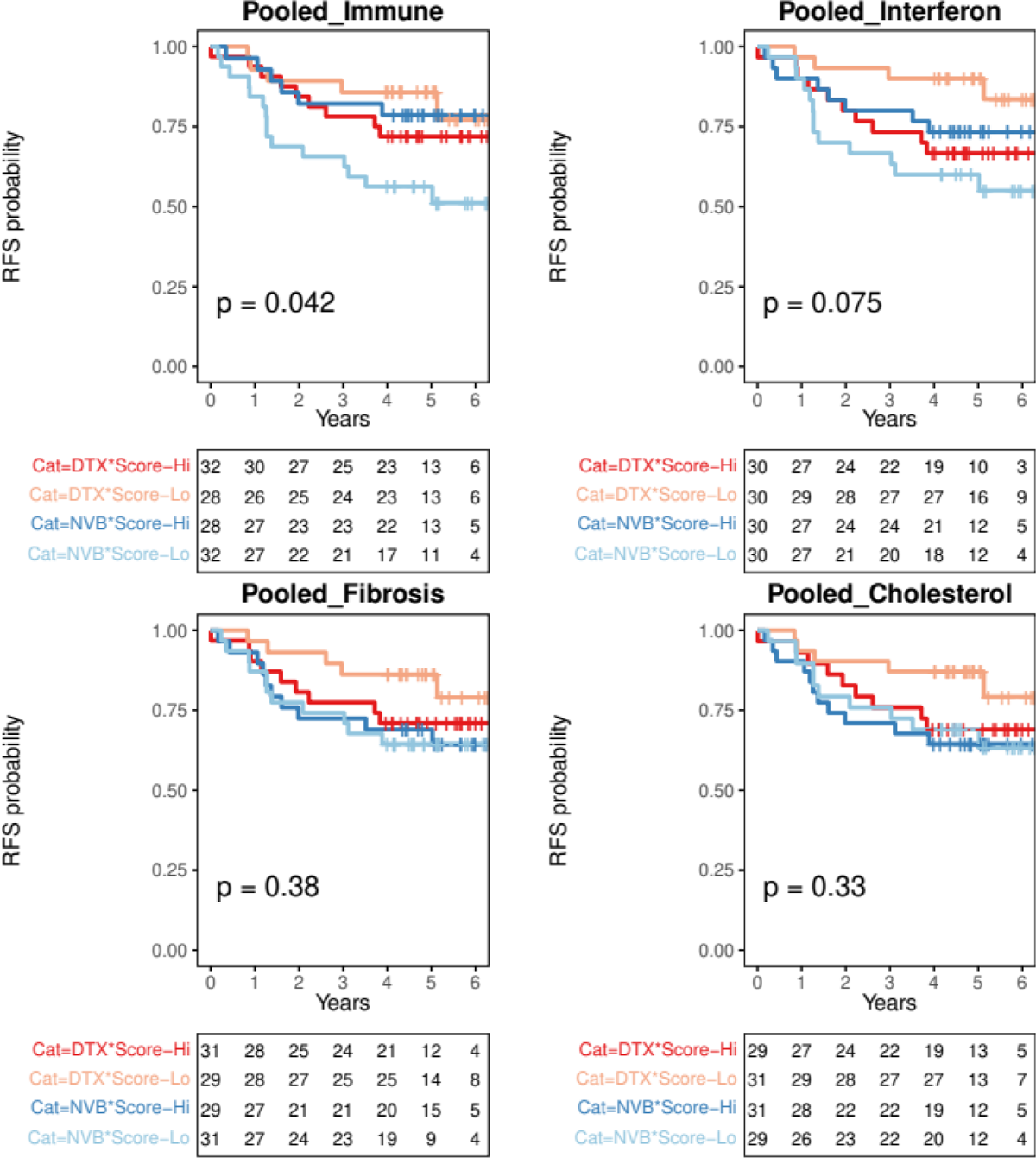
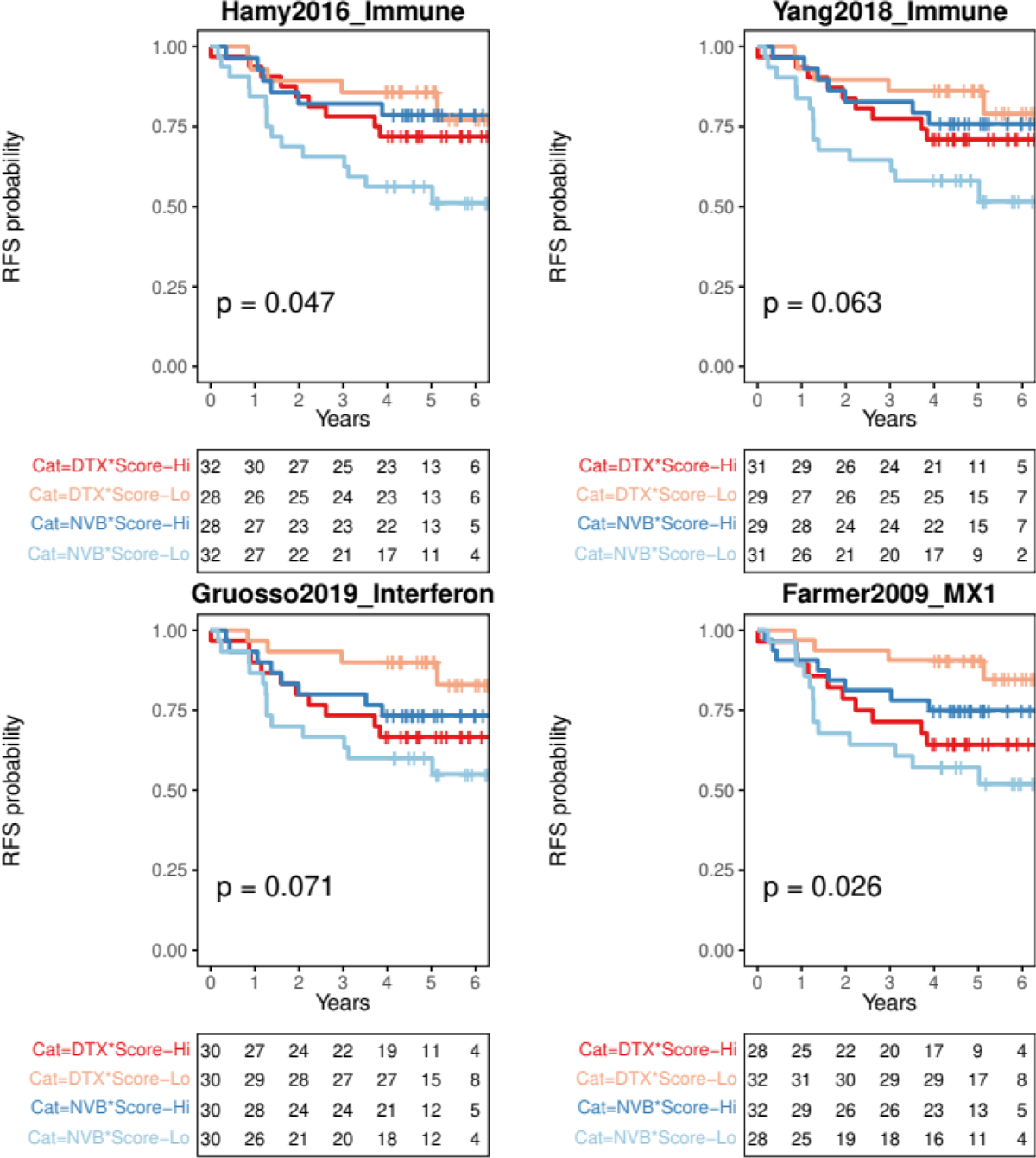
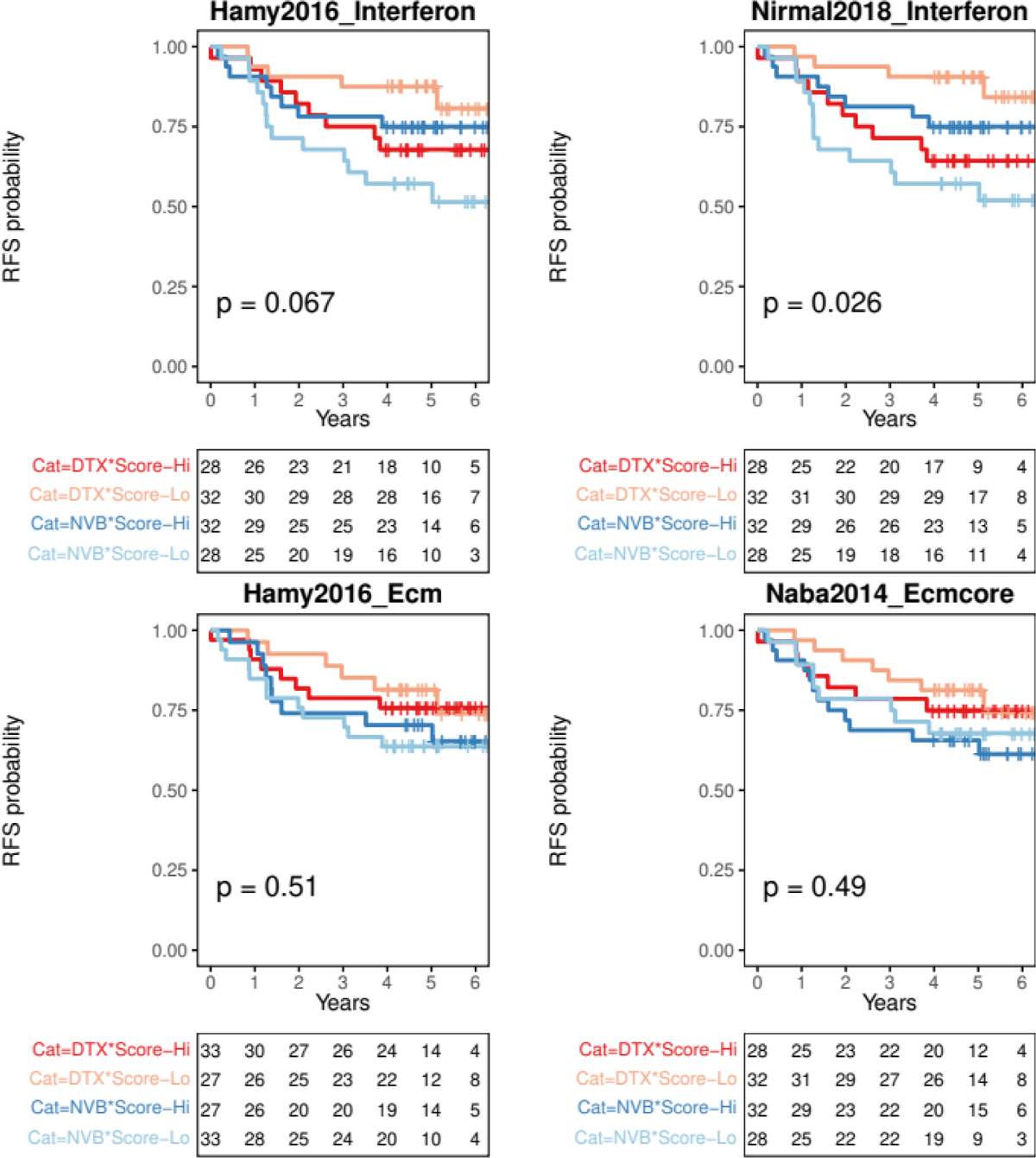
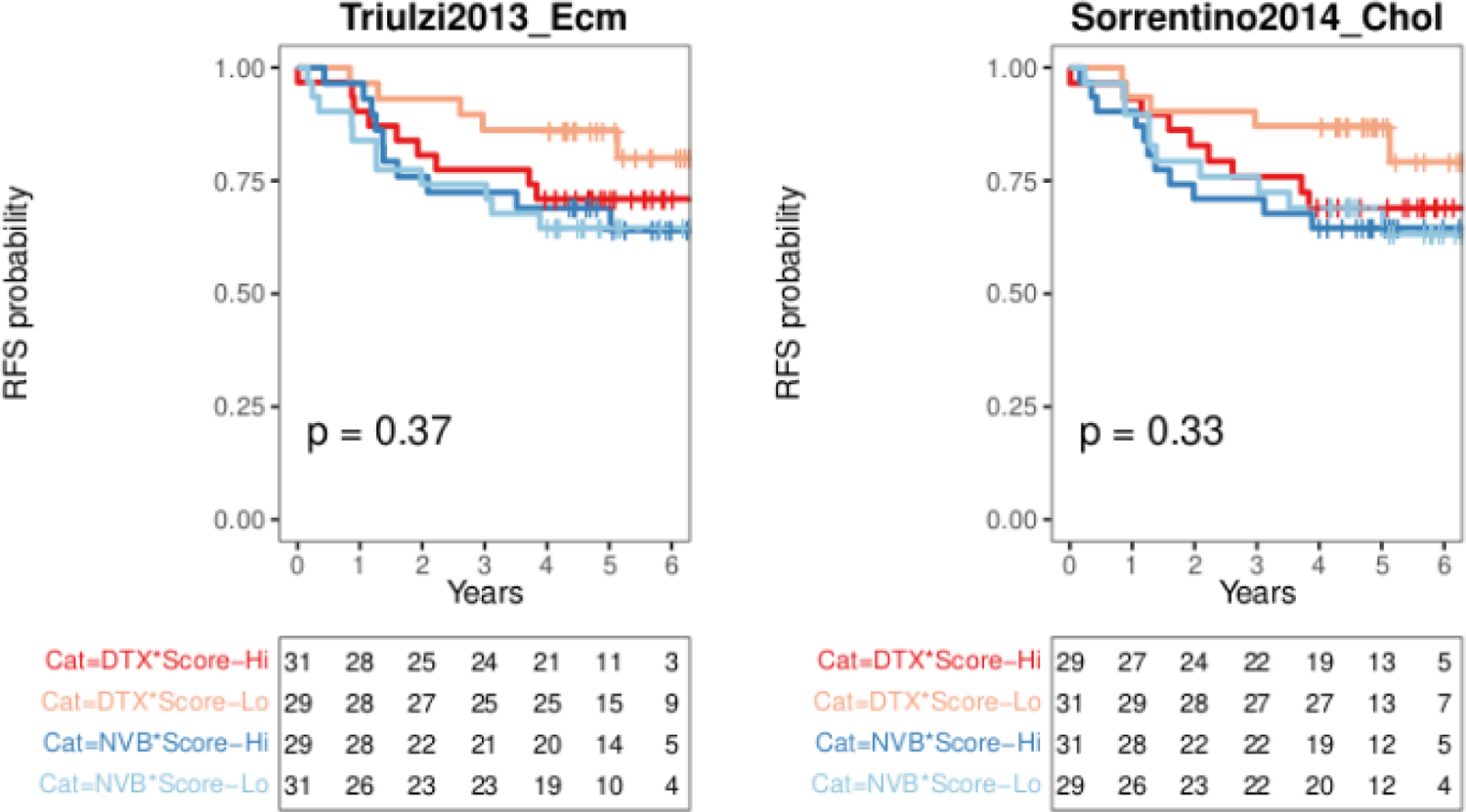
RFS Kaplan-Meyer plots reveal the interaction between MTAs and immune response measures in adjuvant FinHER TNBC. Dark and light red represent Docetaxel with high and low scores respectively. Dark and light blue represent Vinorelbine with high and low scores respectively. High/low classification is based on corresponding medians from TNBC. The median score for TIL (TIL-H&E) in TNBC is 25%. The P-value is computed using the Log-rank test. The table below each Kaplan-Meyer plot shows the number of patients at risk at each time point for each stratum. DTX: Docetaxel - stabilizing MTA. NVB: Vinorelbine - destabilizing MTA.

**Supplementary Figure 14:**
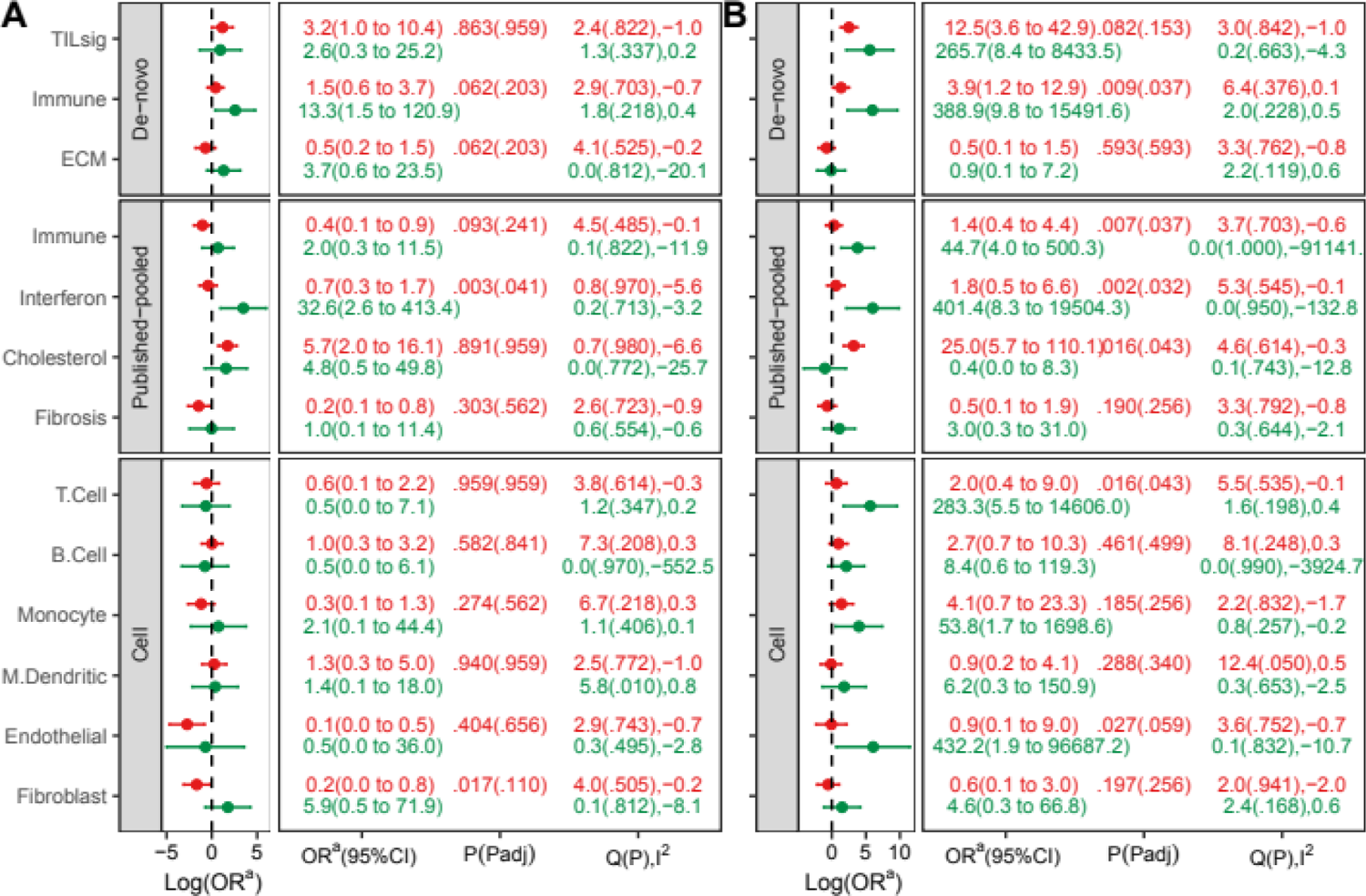
pCR interaction between MTAs and GEO module subsets in the integrated GEO neoadjuvant dataset. (A) TNBC and (B) HR+BC. A positive log(OR) (i.e. OR>1) indicates a higher pCR rate, while a negative log(OR) (i.e. OR<1) indicates a lower pCR rate with an increase in module score in respective treatment regimens. The degree of dataset heterogeneity in each arm is indicated by Cochran’s Q statistic and I^2^ statistic. Q(P),I^2^ represent respectively Cochran’s Q statistic (P-value associated with Q statistic estimated using resampling method) and I^2^ statistic. Red color represents the AAA + Taxane (stabilizing-MTA) regimen, and green color represents the AAA (no-MTA) regimen. ^a^ Odds Ratio.

**Supplementary Figure 15:**
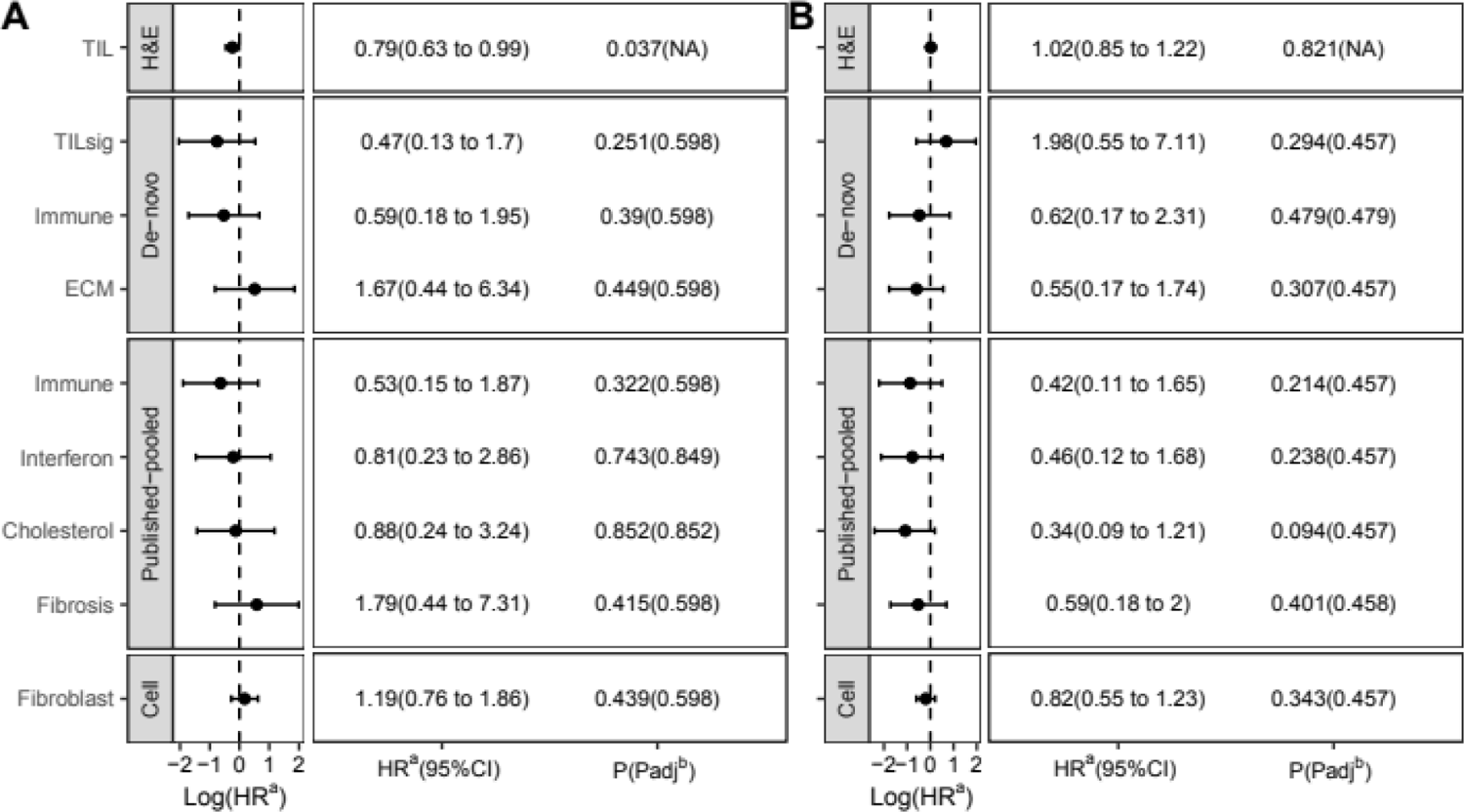
DDFS associations of immune response measures (TIL-H&E/FinGEO module subsets) in the FinHER adjuvant dataset. A positive log(HR) (i.e. HR>1) indicates a higher distant disease recurrence rate, while a negative log(HR) (i.e. HR<1) indicates a lower distant disease recurrence rate with an increase in module score. (A) TNBC; (B) HER2+BC. ^a^ Hazard ratio. ^b^ For FDR correction, the P-value of the DDFS - TIL-H&E association is not considered.

**Supplementary Figure 16:**
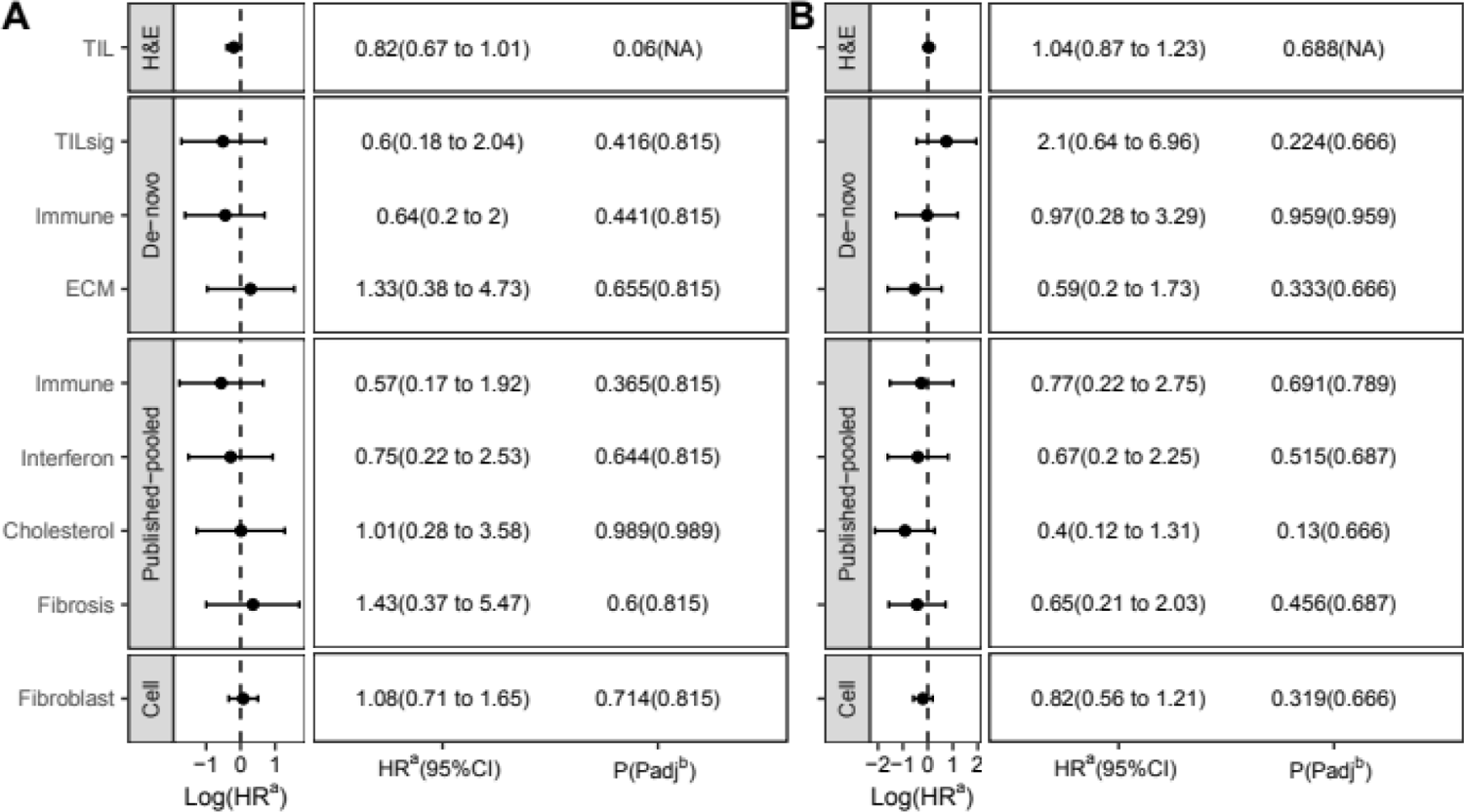
RFS associations of immune response measures (TIL-H&E/FinGEO module subsets) in the FinHER adjuvant dataset. (A) TNBC; (B) HER2+BC. A positive log(HR) (i.e. HR>1) indicates a higher disease recurrence rate, while a negative log(HR) (i.e. HR<1) indicates a lower disease recurrence rate with an increase in module score. ^a^ Hazard ratio. ^b^ For FDR correction, the P-value of the RFS - TIL-H&E association is not considered.

**Supplementary Figure 17:**
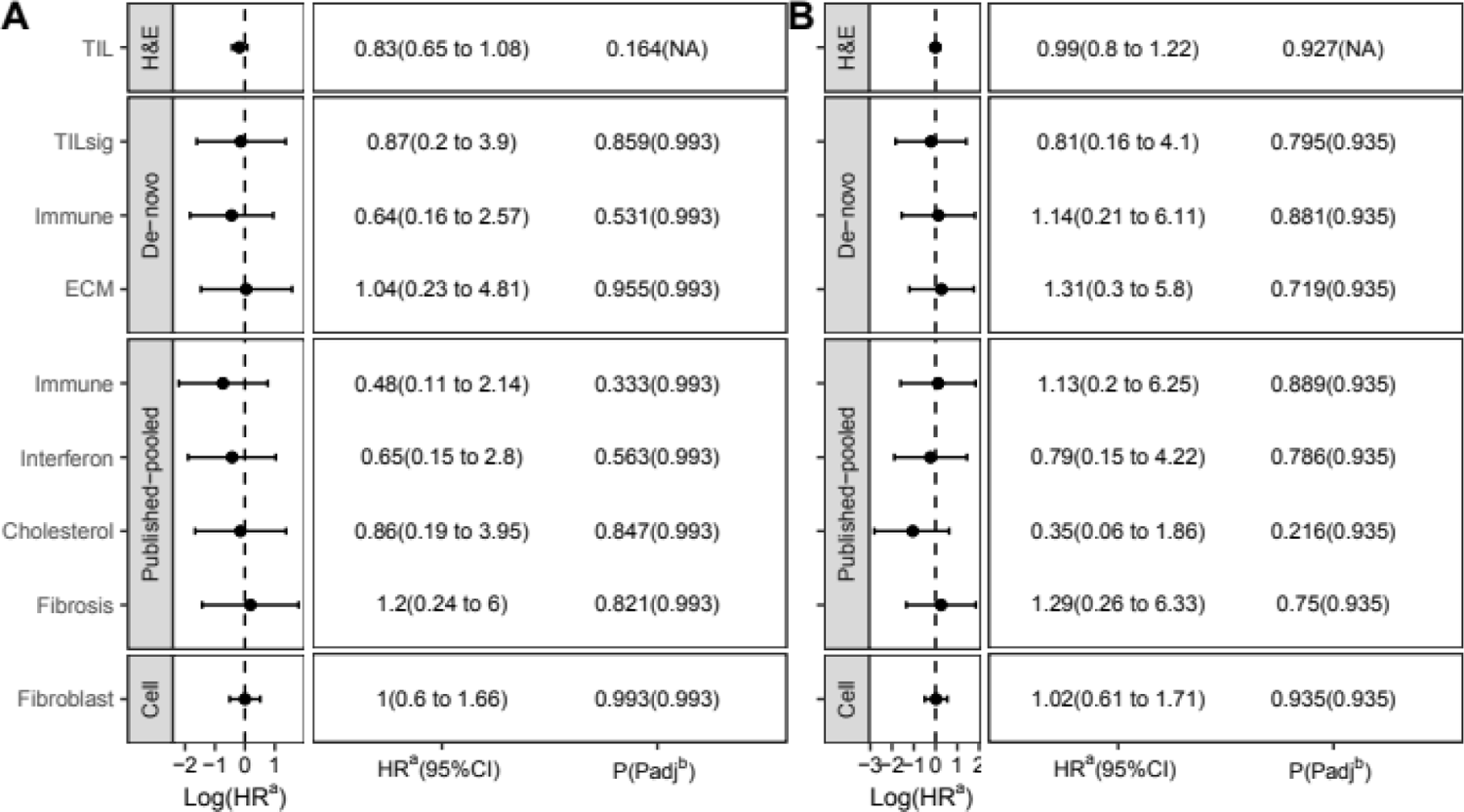
OS associations of immune response measures (TIL-H&E/FinGEO module subsets) in the FinHER adjuvant dataset. (A) TNBC; (B) HER2+BC. A positive log(HR) (i.e. HR>1) indicates a higher death rate, while a negative log(HR) (i.e. HR<1) indicates a lower death rate with an increase in module score. ^a^ Hazard ratio. ^b^ For FDR correction, the P-value of the OS - TIL-H&E association is not considered.

**Supplementary Figure 18:**
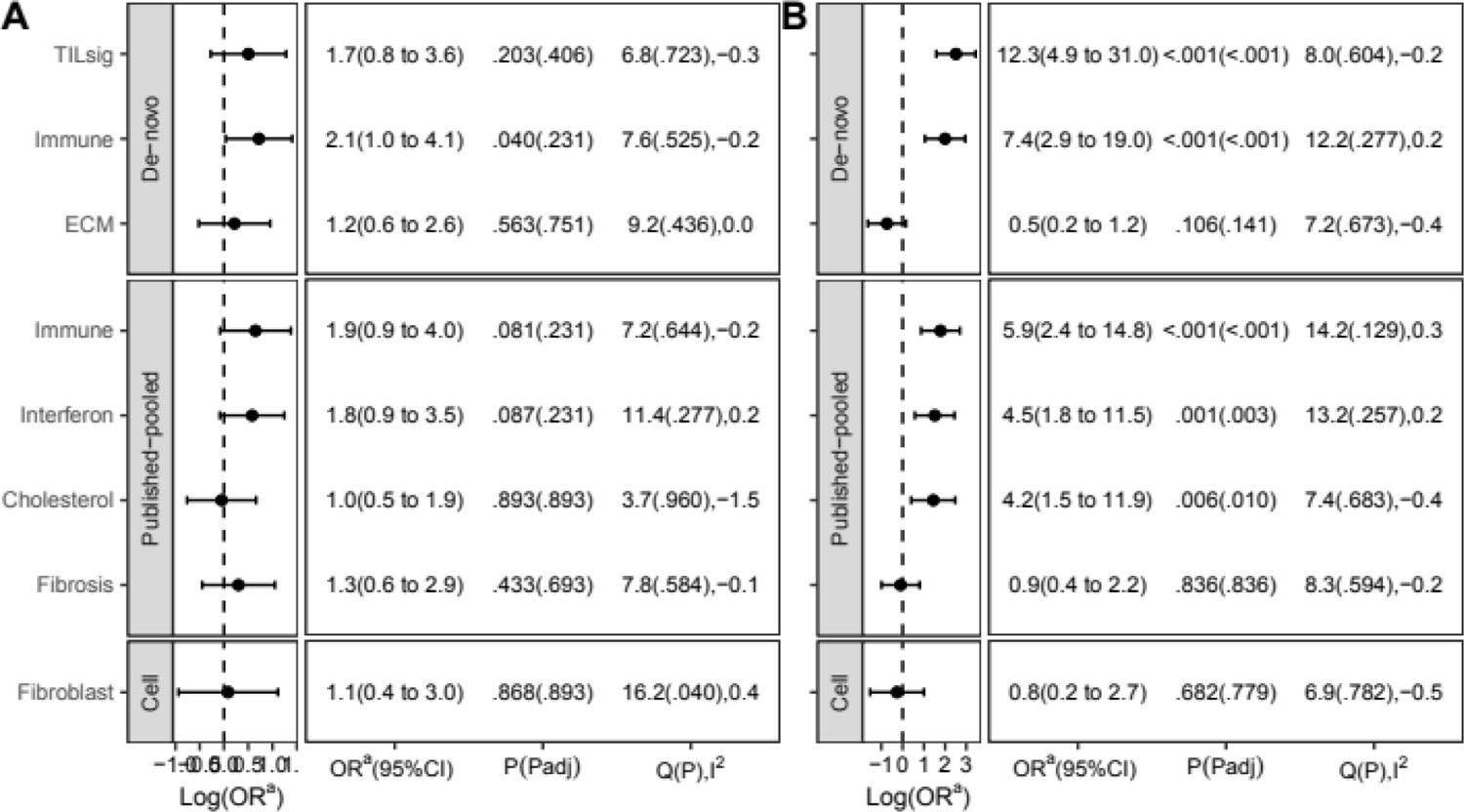
pCR associations of FinGEO module subsets in the integrated GEO neoadjuvant dataset. (A) TNBC and (B) HR+BC. A positive log(OR) (i.e. OR>1) indicates a higher pCR rate, while a negative log(OR) (i.e. OR<1) indicates a lower pCR rate with an increase in module score. The degree of dataset heterogeneity is indicated by Cochran’s Q statistic and I^2^ statistic. Q(P),I^2^ represent respectively Cochran’s Q statistic (P-value associated with Q statistic estimated using resampling method) and I^2^ statistic. ^a^ Odds ratio.

**Supplementary Figure 19:**
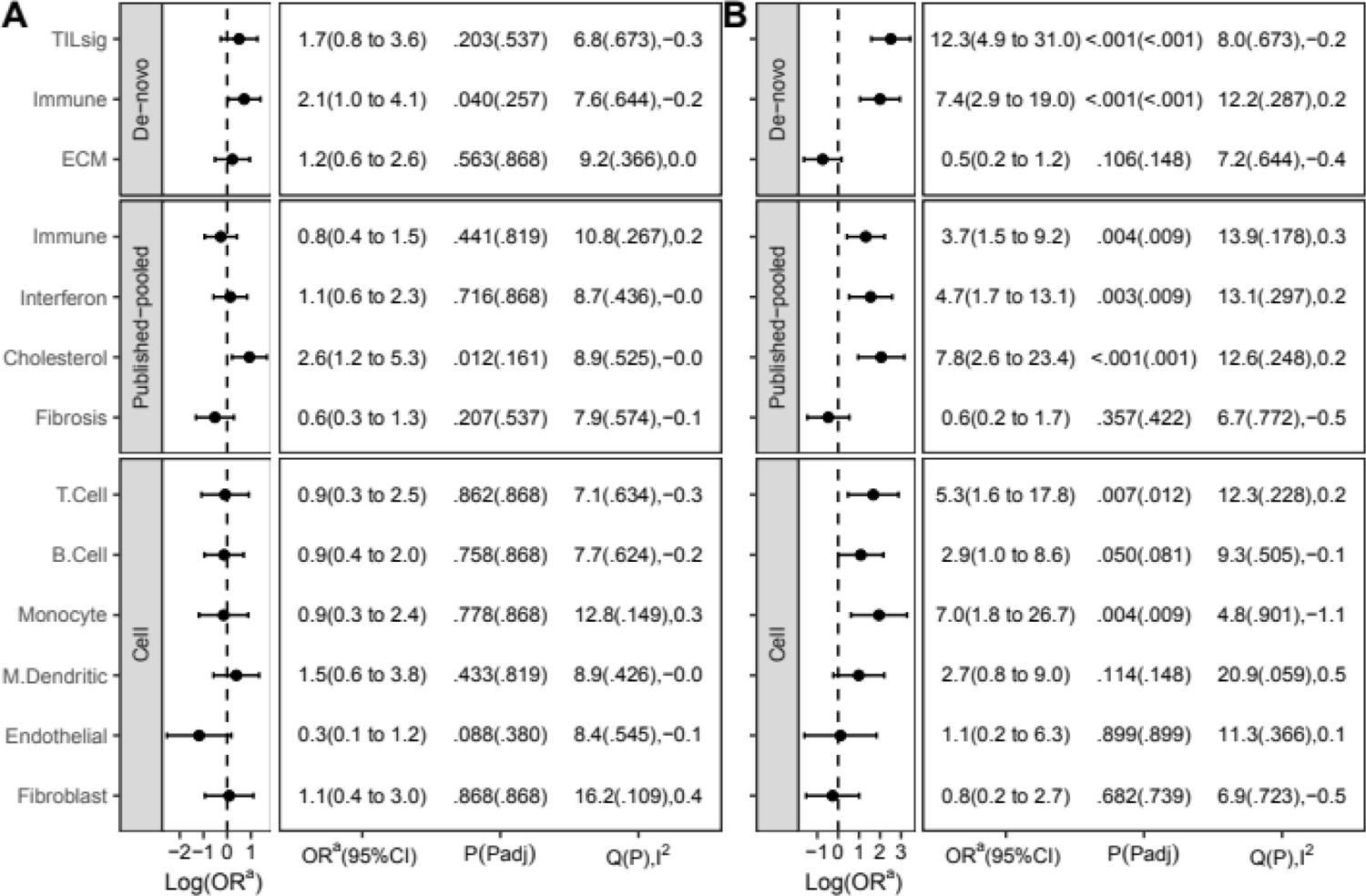
pCR associations of GEO module subsets in the integrated GEO neoadjuvant dataset. (A) TNBC and (B) HR+BC. A positive log(OR) (i.e. OR>1) indicates a higher pCR rate, while a negative log(OR) (i.e. OR<1) indicates a lower pCR rate with an increase in module score. The degree of dataset heterogeneity is indicated by Cochran’s Q statistic and I^2^ statistic. Q(P),I^2^ represent respectively Cochran’s Q statistic (P-value associated with Q statistic estimated using resampling method) and I^2^ statistic. ^a^ Odds ratio.

