## Supplementary Material for "Microtubule targeting agents influence the clinical benefit of immune response in early breast cancer"

### Supplementary file 1

Supplementary to the research article titled "*Microtubule targeting agents influence the clinical benefit of immune response in early breast cancer*"

### Authors and affiliations

Vinu Jose[1], David Venet[1], Françoise Rothé[1], Samira Majjaj[1], Delphine Vincent[1], Laurence Buisseret[1,2], Roberto Salgado[1,3,4], Nicolas Sirtaine[5,6], Stefan Michiels[1,7], Sherene Loi[1,4,8], Heikki Joensuu[9], Christos Sotiriou[1,2*]

1. JC Heuson Breast Cancer Translational Research Laboratory (BCTL), Institut Jules Bordet - Université Libre de Bruxelles, Rue Meylemeersch 90, 1070 Anderlecht, Belgium.
2. Department of Medical Oncology, Institut Jules Bordet - Université Libre de Bruxelles, Rue Meylemeersch 90, 1070 Anderlecht, Belgium
3. Department of Pathology, GZA-ZNA Hospitals, Sint-Vincentiusstraat 20, 2018 Antwerp, Belgium.
4. Division of Cancer Research, Peter MacCallum Cancer Centre, Melbourne, VIC, Australia.

5. Department of Pathology, Institut Jules Bordet - Université Libre de Bruxelles, Rue Meylemeersch 90, 1070 Anderlecht, Belgium.
6. Department of Pathology, Unité Jolimont asbl, Rue Ferrer 159, 7100 Haine-Saint-Paul, Belgium
7. Service de Biostatistique et d'Epidémiologie, Gustave Roussy, Oncostat U1018, Inserm, Paris-Saclay University, labeled Ligue Contre le Cancer, Villejuif, France.
8. Sir Peter MacCallum Cancer Department of Oncology, University of Melbourne, Melbourne, VIC, Australia
9. Helsinki University Hospital and University of Helsinki, Helsinki, Finland

### Table of contents

1. Supplementary materials and methods
2. Supplementary results
3. Bibliography

### 1. Supplementary materials and methods

### TCGA and Metabric datasets

TCGA (1) and Metabric (2) datasets, used for gene module reliability tests, are extracted from the R package MetaGxBreast (3).

### Immune response measurements

#### Gene modules

##### Denovo gene modules

The DAVID functional annotation tool (gene ontology over-representation) uses a modified version of the Fisher exact test (EASE score) to identify over-represented gene ontology (biological-processes) terms in the TILsig gene list compared to the whole list of reliable genes (n=3350) in the array (background gene set) (4–6). The rationale behind using over-represented BP-related gene modules was to reduce noisy genes included in TILsig due to the error associated with the manual counting of TIL from the H&E stained sections by pathologists. Although a genome-wide gene set enrichment analysis is superior to functional over-representation analysis in a set of genes (7), the over-representation analysis is preferred in the present study owing to the limited number of reliable genes present (~ 3000 reliable genes in FinHER). Due to the generation of TIL-BP gene modules from the combined dataset independent of subtypes (see methods), any subtype-specific difference in MTA's interaction with TIL-BP gene modules can be attributed entirely to subtype-specific differences.

#### Published gene modules

The four BPs (immune, fibrosis, interferon, and cholesterol) associated with TIL’s localization, identified by Gruosso et al., are essential to classify TNBC according to TIL’s localization as immune desert, tumor-margin restricted, tumor-stroma restricted, and fully inflamed in the original study (8). We did not classify tumors based on TIL's localization and considered infiltration a gradually progressing process. Further, the cholesterol module, when used independently, cannot be assumed to be explicitly associated with TIL, as its relevance is only in the subgroup of high immune TNBCs with tumor-stroma-restricted infiltration. Therefore, the cholesterol module score from bulk tumor expression profiles can be considered an estimate of general cholesterol signaling not specific to TIL. Identification of cholesterol signaling by Gruosso et al. depends solely on TIL's localization information determined by CD8 immunohistochemistry. Moreover, the de-novo gene modules derived from FinHER did not identify cholesterol signaling as associated with TIL (see methods). Hence, the cholesterol modules considered here measure general cholesterol signaling, and their inclusion in the analysis is solely for completeness.

The original gene module extraction details are available in Supplementary Table 2. The original gene ids are mapped to the NCBI gene id (9) using bioDBnet (https://biodbnet-abcc.ncifcrf.gov/db/db2db.php) (10) if the original id type is not the NCBI gene id. For original gene ids with multiple NCBI gene id mapping, only the stable (in the present study, stable id is the oldest or lowest integer id number) NCBI gene id is considered. The cleaned NCBI gene ids are mapped to the HUGO gene symbol (11) using bioDBnet. The coefficient associated with each original gene id represents each gene's weight and direction of association with the phenotype that the gene module represents. The coefficient is imputed from the respective publications for gene modules without any coefficient specifications. Notably, irrespective of the original coefficient/weight, all genes' weight is set to one to compute the module score while keeping the direction of association intact. The gene modules extracted are available in Supplementary Tables 2.

#### Estimation of immune cells

MCPcounter characterized tumor immune microenvironment (12). Cellular characterization through MCPcounter uses marker gene module scores. The limited number of genes in the FinHER and pooled GEO dataset (3350 and 9184, respectively) may impact MCPcounter reliability as the limited number of genes in marker gene modules compromises its specificity. The reliability of the cell marker gene modules used by MCPcounter is assessed by the method detailed in the main article. The cell marker gene-module statistics, along with the original gene-module to gene module subset correlation, are given in Supplementary Table 2.

#### Statistical analysis

The Rstudio IDE (13) was used to run all analyses in R statistical software (14). The following R packages were used: tidyverse (15), MCPcounter (12), altmeta (16), genefu (17), survival (18), and survminer (19).

##### Immune response's association with clinical response

In the adjuvant FinHER TNBC and HER2+BC datasets, the immune response's association with the clinical response is assessed using stratified univariate Cox regression modeling with strata based on hormone status and treatment regimen. Cox regression modelling formula template used in the adjuvant FinHER dataset: "Surv(time, event) ~ module_score + strata(hormone status + treatment regimen including MTAs)".

In the same manner, in the GEO neoadjuvant TNBC and HR+BC datasets, the immune response's association with the clinical response is assessed using univariate logistic regression modelling after accounting for treatment regimen and dataset id. Logistic regression modelling formula template used in the neoadjuvant GEO dataset: "pCR_status ~ module_score + treatment regimen including MTAs + dataset_id".

##### Interaction between immune response and MTA agents

In the adjuvant FinHER TNBC and HER2+BC datasets, the immune response's interaction with MTA agents on clinical response is assessed using stratified Cox interaction regression modeling with strata based on hormone status and treatment regimen other than MTAs. Cox interaction modelling formula template used in FinHER dataset: “Surv(time, event) ~ module_score * MTA_agent + strata(hormone status + treatment regimen other than MTAs)”.

Similarly, in the neoadjuvant GEO TNBC and HoR+BC datasets, immune response’s interaction with MTA agents on clinical response is assessed using logistic regression modeling after accounting for treatment regimens other than MTAs, and dataset id. Logistic regression modelling formula template used in the neoadjuvant GEO dataset: “pCR_status ~ module_score * MTA_agent + treatment regimen including MTAs + dataset_id”.

#### Fixing PH failures in Cox models

All cox models used in the present study satisfied proportional hazard (PH) assumption criteria, tested with Schoenfeld residual test ($p>0.05$), except for the MTA interaction model with de-novo_TILsig on RFS. PH is fixed by splitting the time to event data into three parts using the cutoffs of 1.43 and 3 years and introducing a three-way interaction term in the cox interaction model (module_score * MTA_agent * time_interval_id). The PH fixed model did not show any significant interaction; hence the estimates from the PH failed model were considered in the analysis for consistency.

### 2. Supplementary results

#### Immune response's association with clinical response in the adjuvant FinHER and neoadjuvant GEO breast cancer datasets

##### TNBC

Contrary to published studies (20–22), no gene modules or cell type estimates used to measure immune response are associated with clinical response in adjuvant and neoadjuvant TNBC (Supplementary Figures 15-19). Note that reliable immune cell type estimates are only available in the neoadjuvant GEO dataset, not in the adjuvant FinHER dataset (see methods). The association  of TIL-H&E with improved DDFS (not on OS/RFS) in the adjuvant FinHER TNBC dataset was reported earlier (Supplementary Figures 15-17) (22).

##### HER2+BC

No measures of immune response were associated with clinical response in adjuvant HER2+BC (Supplementary Figures 15-17). The insignificant association of TIL-H&E with clinical benefit in FinHER HER2+BC was reported previously (22). Notably, the clinical relevance of immune response in adjuvant HER2+BC remains controversial in the published literature(20,22–25). No reliable immune cell types were available in the adjuvant FinHER dataset; only gene modules and TIL-H&E were considered (see methods).

### HR+BC

In neoadjuvant HR+BC, gene modules and cell type estimates used to measure immune response were significantly associated with clinical response (Supplementary Figures 18 and 19), supporting Denkert et al.'s observations (21).
